# Handheld ISFET Lab-on-Chip detection of YAP1 nucleic acid and AR-FL and AR-V7 mRNA from Liquid Biopsies for Prostate Cancer Prognosis

**DOI:** 10.1101/2024.12.30.24317910

**Authors:** Joseph Broomfield, Melpomeni Kalofonou, Costanza Gulli, Sue M. Powell, Rayzel C. Fernandes, Damien A. Leach, Nicolas Moser, Naveed Sarwar, Stephen Mangar, Charlotte L. Bevan, Pantelis Georgiou

## Abstract

Prostate cancer (PCa) is a highly prevalent disease, causing the second largest amount of male cancer deaths worldwide. Currently, the prostate specific antigen (PSA) blood test remains the standard serum prognostic and diagnostic monitoring biomarker but it lacks specificity and sensitivity. PSA testing can lead to invasive biopsies which can result in under detection of clinically significant disease and potential overtreatment of indolent disease. There are promising circulating biomarkers which could facilitate less invasive and more accurate tests, but present challenges in robust quantitation and deployment in clinical settings. This work presents the rapid detection of circulating YAP1 nucleic acid, androgen receptor (AR-FL) and AR-V7 mRNA for PCa prognostics in blood plasma from PCa patients. Sensitive detection of circulating YAP1 nucleic acid, AR-FL and AR-V7 mRNA extracted from PCa clinical samples was achieved with a reverse transcription loop-mediated isothermal amplification (RT-LAMP) assay. Optimisation of mRNA extraction methodologies for reliable detection of circulating mRNA for RT-LAMP and RT-qPCR detection took place. Multiplex testing of circulating AR-FL mRNA and YAP1 nucleic acid on an ISFET Lab-on-Chip platform was readily achieved with bio-electronic signal detection taking place within 15 min. Detection of AR-V7 and AR-FL mRNA could also be achieved simultaneously with the handheld device. Evaluation of clinical data indicated that circulating YAP1 nucleic acid presence in extracted RNA from the blood plasma of patients correlated with more advanced clinical cancer staging (*p* = 0.043) and PSA at diagnosis (*p* = 0.035). The work presents potential for Point-of-Care detection of circulating mRNA from clinical samples for PCa prognostics.

- Circulating YAP1 nucleic acid and AR-FL and AR-V7 circulating mRNA were detected in the blood of prostate cancer patients within 15 min using a RT-LAMP and ISFET handheld biosensor.
- Circulating YAP1 nucleic acid presence was associated with more advanced prostate cancer stage and PSA at diagnosis when detected with the ISFET biosensor. AR-FL and AR-V7 mRNA were only detected in high-risk or metastatic prostate cancer samples.
- This work presents, to the authors’ knowledge, the first endogenous detection of circulating nucleic acids with an ISFET biosensor integrated onto a Lab-on-Chip device for cancer prognostics.

## 1. Introduction

In prostate cancer (PCa) clinics and laboratories, circulating serum concentration of prostate specific antigen (PSA) is the most deployed biomarker. The 33 kilodalton glycoprotein of the KLK3 gene is produced in prostate epithelial cells and is routinely measured in the blood of patients considered at risk of the disease National Institute of Health and Care Excellence (2019). An abnormal PSA result is prostate specific and indicative of abnormal growth but it is not a PCa specific biomarker. As such, elevation of PSA in serum does not preclude benign prostatic hyperplasia presence or urinary tract infections Tchetgen and Oesterling (1997); Zackrisson et al. (2003). Correspondingly, PSA testing often results in the presence of false positives in up to 47 % of real-world cases with the result that invasive treatment is often provided unnecessarily to patients without PCa Martin et al. (2018); Loeb et al. (2014); Lumbreras et al. (2023). PSA testing is additionally utilised to monitor disease and indicate tumour response to treatment. Alternative monitoring technologies directly at point-of-care (PoC), could provide faster clinical decision making and rapid adjustment of treatment options.

More recently, multi-parametric MRI (mpMRI) scanning coupled with PSA testing has seen greater reliability of successfully diagnosing PCa Moore et al. (2023). mpMRI is now recommended after PSA testing and before more invasive biopsies for patients with suspected localised PCa Moore et al. (2023); National Institute of Health and Care Excellence (2019); Conford et al. (2024). However, even when mpMRI is combined with PSA density scores in patients with equivocal radiological features, clinically significant PCa will remain undetected in up to 30 % of cases Arafa et al. (2023); Stavrinides et al. (2021). Prostate biopsies, either trans-rectal ultrasound-guided biopsies (TRUS) or transperineal are often required to gain histological confirmation of PCa as suggested by high PSAs or mpMRI. These biopsy samples undergo immunohistochemical analysis for Gleason Score determination to inform on tumour differentiation and evidence of metastasis. Biopsies however, are invasive and often associated with patients experiencing blood in the semen and urine. Additionally, a small risk of sepsis (2-4 %) and subsequent development of acute urinary retention can occur post-biopsy Sandberg et al. (2024); Bratt (2022). Detection of relevant biomarkers in urine and blood has seen extensive interest in order to remove the necessity of painful procedures for PCa diagnostics and prognostics Nimir et al. (2019); Antonarakis et al. (2017); Reig et al. (2016). Introduction of point-of-care (PoC) biomarker detection in liquid biopsies could result in faster, minimally-invasive testing which can take place directly within the clinic Alexandrou et al. (2023).

Androgens (largely testosterone and dihydrotestosterone) drive differentiation and function of many aspects of the male reproductive system, and the androgen receptor (AR) axis is a major component of prostatic signalling in healthy tissue Davey and Grossmann (2016). However, AR signalling is often dysregulated in PCa and erroneously drives cancer cell proliferation, hence the mainstay of therapy is to reduce androgens and/or directly target the AR Estébanez-Perpiñá et al. (2021). Previous literature has indicated that a greater amount of full-length AR (AR-FL) mRNA in circulating tumour cells (CTCs) is observed in metastatic disease that is resistant to AR-targeting therapy, termed castration-resistant PCa (mCRPC) compared to hormone sensitive PCa (HSPC) patients Nimir et al. (2019). AR gene gain in circulating exosomes and greater AR mRNA expression in circulating tumour cells (CTCs) from mCRPC patients are associated with reduced progression-free survival (PFS) and overall survival (OS) following treatment with AR-targeting abiraterone or enzalutamide Del Re et al. (2021); Silberstein et al. (2017). In each instance, increased AR circulating cell-free DNA (ccfDNA) or mRNA levels were also associated with increased likelihood of detection of a variant of AR termed AR-V7 Del Re et al. (2021); Silberstein et al. (2017). Expression of AR variants (AR-Vs) can be selected for in response to AR-targeting therapies, creating a more aggressive disease that is no longer responsive to therapy. These AR-Vs often involved truncation of the AR protein to exclude the ligand binding domain region, resulting in a constitutively active protein which is targeted by all current anti-AR therapies. AR-V7 is a common variant of the AR and its detection in circulation correlates with reduced PFS and OS in patients treated with enzalutamide and abiraterone Antonarakis et al. (2014, 2017); Aasems et al. (2021).

YAP1 is another potential prognostic biomarker as its over-expression in PCa tissue is associated with worse diseasefree survival and OS in various cancer types Sun et al. (2015). YAP1 is a mechanosensory protein, able to transmit cues from the extracellular environment to the nucleus and chromatin to drive processes associated with invasion and metastasis and as such is associated with cancer progression.Sun et al. (2019); Baek et al. (2022); Hu et al. (2022). Previous work has indicated that YAP1 expression in healthy tissue is lost in low grade PCa Cheng et al. (2020). However, in high grade PCa, YAP1 expression is upregulated. This distinction of YAP1 nucleic acid expression between low grade and high grade could provide a quantitative circulating biomarker for PCa progression. Gene copy numbers of YAP1 are additionally increased at metastatic sites when compared to matched primary tumours, indicating the relevance of both YAP1 DNA and mRNA for potential prognostic biomarkers Collak et al. (2020).

YAP1 is a positive regulator of the AR in PCa cell lines which mimic hormone naive PCa Kuser-Abali et al. (2015). In VCaPs, a PCa cell line, ERG-mediated invasion is dependent on YAP1 function Nguyen et al. (2015). Since the VCaP cell line contains the constitutively active TMPRSS2-ERG gene fusion, YAP1 is implicated with another mRNA biomarker associated with more advanced clinical staging and high Gleason Scoring Hägglöf et al. (2014). Contrastingly, it has been observed that YAP1 is targeted by a microRNA miR-375, which is upregulated in metastatic PC Selth et al. (2017). This indicates that YAP1 could potentially act as a temporal mRNA marker, which is upregulated in high grade HSPC and downregulated in mCRPC Selth et al. (2017). While circulating miR-375 presence positively correlates with CTC count, YAP1 circulating mRNA or DNA presence has not previously been explored in cancer. Selth et al. (2017)

While the PSA test detects protein concentrations, nucleic acid biomarkers in the blood including ccfDNA, cfRNA and miRNAs can additionally provide valuable information on PCa prognostics. Detection of relevant nucleic acid sequences is often achieved with nucleic acid amplification tests (NAATs). The quantitative reverse transcription polymerase chain reaction (RT-qPCR) is a robust and simple molecular biology technique capable of sensitive and specific detection of mRNA sequences. For these reasons, RT-qPCR is typically considered the ‘gold standard’ of NAATs. More recently digital droplet qPCR (ddPCR) has been utilised for ultra-sensitive and absolute quantification of circulating nucleic acids, albeit at the trade-off of greater complexity Danila et al. (2016); Stitz et al. (2024). However, both ddPCR and RT-qPCR lack PoC compatibility since the cycling between dsDNA separation (90 - 95 °C) and primer annealing (55 - 65 °C) in these techniques requires specialised and expensive thermal cycling equipment Becherer et al. (2020). Therefore, investigation of the usage of isothermal nucleic acid amplification tests (NAATs) could present utility for PoC diagnostics and prognostics Rodriguez-Manzano et al. (2021); Koo et al. (2016); Wang et al. (2020); Broomfield et al. (2022b). Utilisation of inexpensive equipment, as well as faster time to positive (TTP) results which could be introduced directly within clinics supports the viability of isothermal NAAT for PoC testing Becherer et al. (2020); Song et al. (2022); Rodriguez-Manzano et al. (2016). For example, reverse transcriptase loop-mediated isothermal amplification (RT-LAMP) is a highly specific and rapid technique for RNA sequence detection Notomi et al. (2000). LAMP was originally introduced by Notomi *et al* 2000, and updated by Nagamine *et al* 2002, as an isothermal and exponential amplification method Notomi et al. (2000); Nagamine et al. (2002). TTPs for LAMP can range from 5 - 60 min, and the technique requires 4 to 6 primers, typically targeting a larger region of DNA than qPCR primers Notomi et al. (2000).

Previous work has established RT-pHLAMP assays are capable of quantitative and sensitive detection of YAP1, AR-V7 and AR-FL mRNA from PCa cell lines within 15 min Broomfield et al. (2022a, 2023). These reactions utilised an augmented RT-LAMP assay without standard buffering agents to enable real-time monitoring of pH change generated as a result of sequence-specific amplification reactions. These assays were successfully implemented onto Lacewing, an ion-sensitive field-effect transistor (ISFET) - based Lab-on-Chip (LoC) system which has the potential to be operated directly at PoC Broomfield et al. (2022a); Moser et al. (2019). Utilising unmodified complementary metaloxide semiconductor technology, this LoC system can result in a rapid PoC device with the potential for inexpensive commercialisation. The standard passivation (sensing layer of the process of the ISFET is capable of detecting the rate of pH change in solution Moser et al. (2016); Broomfield et al. (2024). Previous work using an ISFET biosensing for RT-pHLAMP or pHLAMP detection has demonstrated the capability of rapid SARS-CoV-2 detection Rodriguez-Manzano et al. (2021). Detection of synthetic mutated DNA associated with breast and colorectal cancer progression further indicates the potential for cancer patient monitoring with the ISFET Lab-on-Chip device. Kalofonou et al. (2020); Alexandrou et al. (2020); Mantikas et al. (2023). Equation 1 illustrates the principle of detection due to the elongation of dsDNA during LAMP amplification resulting in the production of a proton per nucleotide addition.

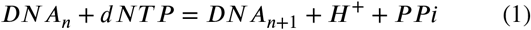

Through the release of protons during an RT-LAMP amplification event, a pH change is induced which can be observed as a rise in voltage on the ISFET LoC.

This work shows potential for the first ISFET LoC device for simultaneous detection of circulating nucleic acid markers for PCa prognosis from liquid biopsies. Detection of AR-FL and AR-V7 mRNA and YAP1 nucleic acid extracted from blood plasma from PCa patients will be explored utilising RT-pHLAMP. The presence of these markers could provide rapid, clinically relevant information at PoC. Figure 1 illustrates the proposed Lacewing ISFET Lab-on-Chip technique for generation of clinically relevant information compared to current PCa prognostics (Gleason Score and PSA) and RT-qPCR.

**Figure 1:**
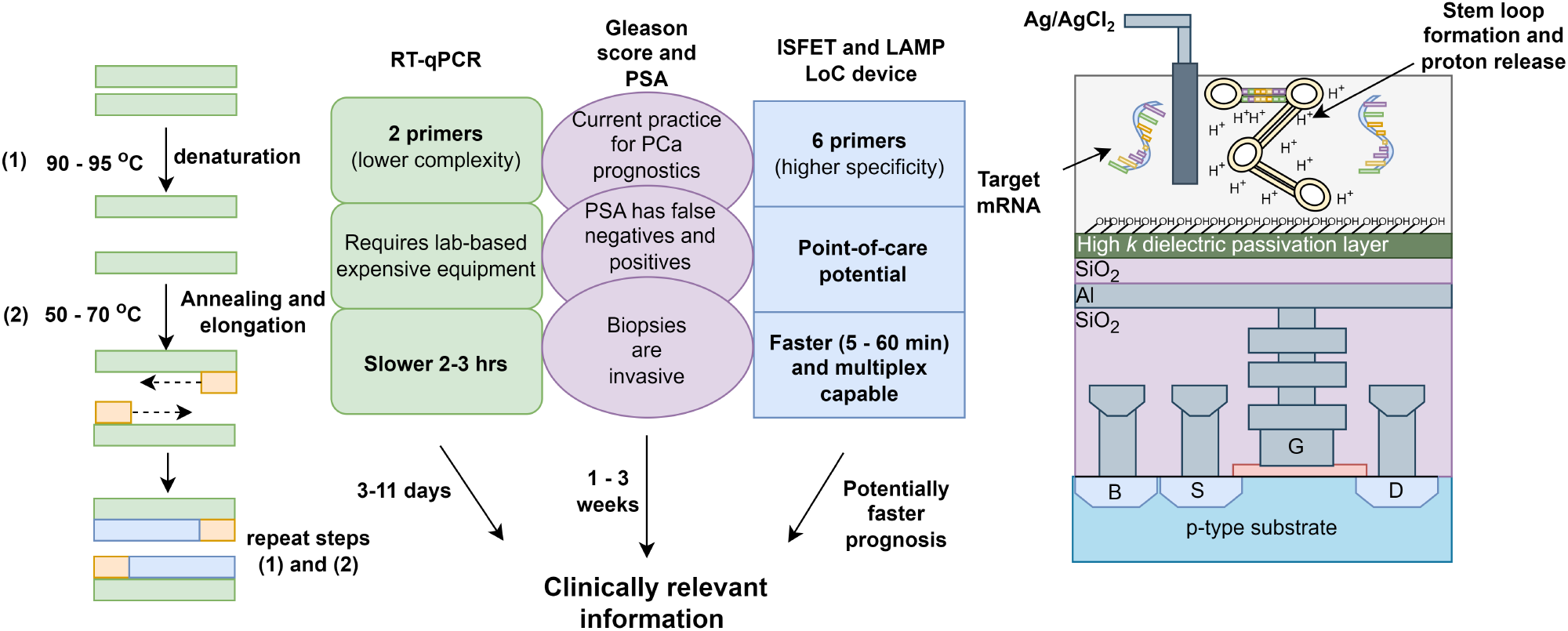
Comparison of techniques for prognostics including RT-qPCR, Gleason Score and PSA and the ISFET Lab-on-Chip device proposed in this work. Time for PCa RT-qPCR results for mRNA from CTCs is between 3-11 days Sieuwerts et al. (2018). Time for results from a TRUS by the NHS is one to three weeks Cancer Research UK (2022).

## 2. Results and discussion

### 2.1. Extraction optimisation for RT-pHLAMP detection

Preliminary testing with RT-pHLAMP assays occurred directly with plasma. Blood plasma spiked with synthetic YAP1 RNA has previously resulted in improved time to positive (TTP) and higher sensitivity (100 copies per reaction) when compared to serum Broomfield et al. (2022a). However, no amplification was observed after 35 minutes from the onset of the reaction in patient samples P1 - P14 (data not shown). Circulating mRNA is a low abundance marker in the blood. Only 1-100 pg / *μ*L of circulating free RNA is present in plasma or serum Norgen Biotek (2022). As such, RNA extraction techniques were utilised for successful RNA detection.

Optimisation of RNA extraction was essential for accurate quantification of circulating mRNA expression. Spiked extractions were initially conducted with a complex system of mixed-male commercial plasma and spiked-in synthetic YAP1 mRNA. Two different extraction methods were attempted using a Norgen Biotek plasma/serum RNA purification mini kit and a standard TRIzol LS extraction. (Supplementary Figure S1). Synthetic RNA was readily detectable down to 30 copies per reaction with the TRIzol LS reagent. The reaction remains relatively quantitative (R^2^ = 0.874) given the extraction of synthetic RNA within a complex medium (plasma) (Supplementary Figure S1 **a**). Detection post Norgen Biotek extraction was possible down to 10^4^ copies, albeit large standard deviations of the RT-pHLAMP reaction reduced the quantitative reliability of the assay. As such, the TRIzol LS reagent extraction was utilised for extraction of PCa plasma samples for YAP1, AR-FL and AR-V7 mRNA. Similarly, quantitative detection of AR-FL mRNA post-extraction with TRIzol LS reagent was observed down to 1000 copies per reaction (R^2^ = 0.964). Detection of a smaller mRNA fragment of 228 base pairs (bp) was achieved more successfully than with a 350 bp fragment. This could be indicative of greater stability of the smaller fragment in plasma. AR-V7 mRNA spiked into commercial plasma additionally could detect down to 100 copies per reaction after TRIzol LS reagent (Supplementary Figure S1 **c**). The AR-V7 assay remained quantitative within the range of 1 x 10^8^ down to 100 copies (R^2^ = 0.962), albeit with some variation between technical repeats. Once it had been determined that the RT-pHLAMP assays were capable of relevant mRNA detection after TRIzol LS RNA extraction, detection in clinical samples took place.

For Trizol LS mRNA extraction, gDNA was not removed. For the YAP1 RT-pHLAMP assay, where the primers do not span exon-exon boundaries, both YAP1 ccfDNA and mRNA could be detected. It was hypothesised that this would increase the incidence of detected YAP1 within the patient sample cohort, potentially culminating in a more robust prognostic test. As such, ‘YAP1 nucleic acid’ is utilised to describe the target of the YAP1 RT-pHLAMP assay. The primers for AR-FL cross exon-exon boundaries and its variant AR-V7 is thought to originate from aberrant mRNA splicing Liu and Dong (2014). As such, the target of these assays are referred to as ‘mRNA’.

To ensure that extractions were reproducible in PCa liquid biopsy samples, P1-P4 EDTA samples were extracted in triplicate and both YAP1 and *β*-actin housekeeping expression were recorded with two-step RT-qPCR. Figure 2 **b** illustrates that the TRIzol extractions were highly reproducible across three *β*-actin repeats, which was crucial to ensure consistent detection of circulating RNA. YAP1 nucleic acid detection was also consistent across the four patient samples by both RT-qPCR and RT-pHLAMP. There was no statistical significance between the two repeats with a two-tailed equal variance unpaired t test, with either the RT-qPCR or RT-pHLAMP reactions. Two repeats were utilised here for YAP1 nucleic acid detection to ensure enough extracted plasma samples were preserved for further testing.

**Figure 2:**
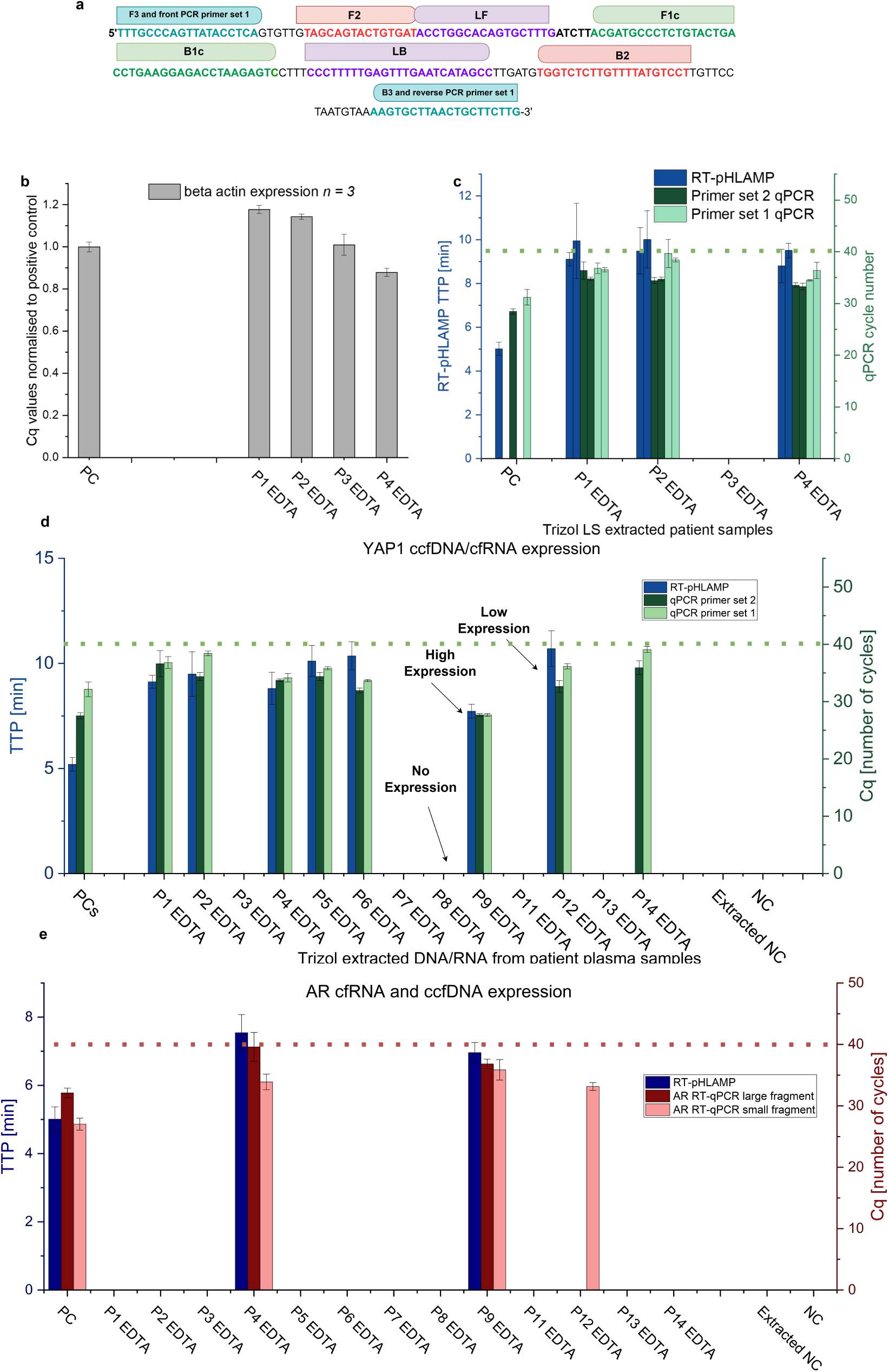
**(a)**: The YAP1 DNA sequence that is targeted by the RT-pHLAMP primers and PCR primer set 1. Primer directions are shown, reverse primers are the reverse complement of the DNA sequence. **b**: *β*-actin RT-qPCR expression across three individual extractions from EDTA plasma across four patient samples. Error bars represent ± one standard deviation. **c** RT-qPCR experiment showing the detection of YAP1 expression across two TRIzol extractions. **d**: The detection of both YAP1 RT-qPCR assays and the developed YAP1 RT-pHLAMP reaction across patient samples P1-P14 (excluding P10). **(e)**: The detection of androgen receptor mRNA from patient samples P1-P14 (excluding P10) with RT-pHLAMP and large and small fragment RT-qPCR. In the graphs the dotted lines represent the PCR cycle cut off of 40 for averaged technical repeats.

Figure 2 illustrates the nucleic acid presence of YAP1 in patient samples P1 - P14. P10 was omitted from this analysis due to insufficient sample volume. RT-pHLAMP detection of YAP1 was compared to the detection of two different RT-qPCR YAP1 fragments. RT-qPCR YAP1 primer set 1 covers the same nucleic acid sequences as the F3 and B3 primers developed for the RT-pHLAMP reaction. This ensured that the specific nucleic acid sequence that the RT-pHLAMP reaction detects is present in the extracted patient samples, particularly since nucleic acids in the blood are often degraded Yao et al. (2020); Qin et al. (2016). For RT-qPCR primer set 1, primer concentrations were optimised to ensure a relevant limit of detection was achieved (Supplementary information Figure 2). RT-qPCR YAP1 primer set 2 targets a separate region of the YAP1 nucleic acid sequence. This primer set confirmed that an alternate region of the YAP1 sequence was present besides the region targeted with RT-pHLAMP and RT-qPCR primer set 1. Figure 2 indicates that the YAP1 RT-pHLAMP benchtop assay can detect YAP1 nucleic acid presence in the same samples as both RT-qPCR assays, with the exception of the sample from patient 14, which exhibited the slowest Cq cycle number for the same region as the RT-pHLAMP primers with 39.08 ± 0.54 cycles. In this instance, it is likely that the limit of detection of RT-pHLAMP reaction is not sufficient for reliable detection at this low concentration of YAP1. YAP1 nucleic acid presence was detected in 62 % of patient samples (*n = 8*) by RT-qPCR, indicating a reasonable prevalence within this patient cohort. Detection with the benchtop RT-pHLAMP reaction rapidly occurred within 12 min for all experiments. In comparison, all PCR experiments were completed within 2 hr 30 min and required conversion of RNA to cDNA before the assay was performed.

The YAP1 RT-pHLAMP benchtop assay was capable of distinguishing high and low concentration detection of YAP1 nucleic acid from TRIzol extracted clinical samples (Figure 2 **d**). Specifically, P9 EDTA extracted RNA exhibited the highest YAP1 nucleic expression across the three NAAT assays. With RT-pHLAMP detection of YAP1 in P9 was observed within 7.72 ± 0.33 min. Other slower TTP values including from P5, P6 and P12 were readily distinguishable from the faster TTP of P9 (*p* = 0.002, < 0.001 and < 0.001 respectively, unpaired t-test). The Pearson’s correlation between the YAP1 RT-qPCR primer set 1 and the RT-pHLAMP assay was 0.504, which indicates a positive trend but a limited correlation. While these assays detect the same nucleic acid fragment, contaminants from extracted patient samples are likely to affect Cq and TTP values with RT-qPCR and RT-pHLAMP reactions differently. In fact, LAMP is often a preferred technique on account of its robust amplification in the presence of traditional qPCR inhibitors, such as bile salts, IgG and urea which are all present in human blood plasma Nwe et al. (2024).

Recent research on digital droplet qPCR has additionally indicated that large proportions or all of HSPC cohorts exhibit basal AR-FL cfRNA expression Nimir et al. (2019); Stitz et al. (2024). This copy number of AR cfRNA expression in HSPC patients has previously been reported in the range of 0-15 copies per ml of plasma Nimir et al. (2019). Since 250 *μ*L of plasma was utilised for RNA extraction in this work, it was anticipated that this basal expression would not be detected in the AR-FL RT-pHLAMP assay. As such, the proposed PoC assay would detect AR-FL expression above the basal expression in HSPC only. This could be indicative of AR-FL amplification which has previously been implicated in reduced PFS and OS of PCa patients Silberstein et al. (2017).

The AR-FL RT-pHLAMP assay previously described was adapted to improve the sensitivity of the reaction to TRIzol extracted mRNA samples (Supplementary Figure S3) Broomfield et al. (2023). It was assumed that the sensitivity of the assay from extracted samples down to 1000 copies per reaction would be insufficient to detect circulating AR-FL mRNA amplification. NaOH, MgSO_4_, BSA and RT-pHLAMP primer concentrations were adjusted to improve the TTP of the reaction and allowed for detection of AR-FL mRNA in patient samples.

Confirmation of presence of AR-FL mRNA was additionally conducted with two separate RT-qPCR experiments. AR-FL RT-qPCR large fragment detected a similar sized sequence as the LAMP reaction and would also only detect mRNA sequences. It was ascertained experimentally that the RT-qPCR small fragment reaction using primers F3 and B3 from RT-pHLAMP was less efficient than using the forward PCR primer sequence from the small fragment RT-qPCR assay, and B3 from the RT-pHLAMP reaction. As such, the more efficacious assay was used for the large fragment RT-qPCR assay. Further explanation of this PCR primer set, including sensitivity testing and primer concentration testing can be found in the Supplementary Information section (Figure S4). The small fragment RT-qPCR assay detected a smaller 50 bp fragment, which did not span exon-exon boundaries. As such, both cfRNA and gDNA could be amplified with this RT-qPCR assay. The small fragment AR-FL RT-qPCR assay can be utilised to observe AR-FL nucleic acid presence after TRIzol LS extraction. Figure 2 indicates that several (*n=3*) samples contained AR-FL nucleic acid presence. However, in P12, amplification was only observed in the assay which detects both RNA and DNA. This indicates that AR-FL mRNA amplification was not present in P12 but AR-FL ccfDNA was detected. Correspondingly, this provides further evidence that the AR-FL RT-pHLAMP assay is specific to AR-FL mRNA amplification detection.

This work indicated that the benchtop AR-FL RT-pHLAMP reaction showed 100 % concordance with the AR-FL large fragment RT-qPCR assay. YAP1 nucleic acid presence was additionally concordant with 12 out of 13 (92.3 %) of the samples with both YAP1 RT-qPCR assays. The sensitivity of these two RT-pHLAMP assay was deemed suitable for testing with the Lab-on-Chip device for point-of-care utility.

#### Lab-on-Chip detection of patient samples

Initial Lab-on-chip detection was optimised with one positive YAP1 nucleic acid sample (P4 EDTA) and one negative sample (P3 EDTA) as previously determined by the RT-qPCR and RT-pHLAMP assays (Supplementary Figure S5). These experiments were conducted in triplicate and utilised single-well manifolds, which do not have multiplex capability. This LoC method has previously been described for YAP1, AR-V7 and AR-FL mRNA detection from PCa cell lines Broomfield et al. (2022a, Broomfield et al. 2023). The biosensor is composed of a 78×56 array of ISFET pixels which individually record the rate of pH range in the RT-pHLAMP solution. The output from the ISFET pixels is averaged and is recorded as the sensor output. Single-well LoC detection was successful in rapidly detecting YAP1 nucleic acid presence in P4 EDTA while P3 EDTA showed little to no deviation from the sensor drift. The presence of the target YAP1 sequence resulted in the production of protons during nucleotide addition onto the double stranded DNA formed during the RT-pHLAMP reaction. During an amplification (positive sample) event, the voltage will rise above the expected sensor drift. After termination of amplification, often several minutes later, the biosensor output returns to sensor drift. An example of a sensor output is recorded in Supplementary Figure S5. Conversion of the sensor output to a pH change has previously been reported Yu et al. (2019); Rodriguez-Manzano et al. (2020). Quantification of DNA and pH change were recorded post-experiment to confirm the results of the sensor. The TTP of YAP1 nucleic acid detection in P4 with the LoC device was observed within 9.63 ± 1.29 min compared to 8.81 ± 0.66 min in the benchtop reaction (Supplementary Figure S5). These TTP values are largely comparable, with the ISFET LoC device showing slightly slower TTPs than the benchtop which has previously been observed Broomfield et al. (2022a, Broomfield et al. 2023).

Once it had been confirmed that the detection of extracted RNA samples was reproducible on the LoC, adaptation to a two-well manifold for multiplex detection transpired. The acrylic manifold and post-processing code were adjusted for dual-well detection on the ISFET sensor. The post-processing code previously used to determine positives and negatives on the sensor was adapted to select only data on one half of the sensor. This allows for partitioning of the two wells for individual analysis. Figure 3 shows the first frame of the ISFET biosensor with the dual-well manifold attached. The two wells on this manifold have a volume of 15 *μ*L instead of 20 *μ*L. The volume of the wells was reduced to increase the size of the acrylic wall between the reaction chambers, limiting potential leakage between the wells. It was not anticipated that the reduced sample size would affect the RT-pHLAMP reaction, since the volume of benchtop RT-pHLAMP reactions was 10 *μ*L. The two wells are easily distinguishable on the ISFET array surface, where yellow pixels are in range for detection of pH change (Figure 3). Lab-on-Chip detection was achieved for the simultaneous detection of AR-FL and YAP1 RT-pHLAMP assays. The data recorded in these experiments is summarised in Figure 3. Figure 3 illustrates three patient samples detected with the device which have different iterations of YAP1 nucleic acid and AR-FL mRNA presence. There were no instances in this cohort where AR-FL mRNA was present but YAP1 nucleic acid was not. Amplification curves for the other patient samples can be found in the Supplementary Information file (Figures S6 and S7). The LoC results were 100 % concordant with the benchtop RT-pHLAMP assay for both YAP1 and AR-FL detection. This indicated that the simultaneous PoC detection of these markers retained the high sensitivity and specificity of the benchtop assays. While the pH output for the device for P4 is lower than the P12 example, observable amplification events from the sensor outputs were easily distinguished. Species in solution can reduce the efficacy of ISFET detection to pH change, which could account for the reduced pH signal observed. As with previous LoC detection, slightly slower TTPs were observed with the point-of-care device relative to the benchtop. For example, benchtop YAP1 and AR-FL RT-pHLAMP assays detected their respective targets within P4 in 8.81 ± 0.77 and 7.55 ± 0.53 min. Lab-on-Chip detection was observed within 10.5 and 7.75 min (Table 1). Supplementary Figure S8 shows the relationship between benchtop and LoC TTP values for each patient sample (Pearson’s coefficient 0.7) indicating a medium to strong positive correlation. AR-FL mRNA detection with RT-pHLAMP was observed in one metastatic patient and one high-risk patient. ccfDNA and cfRNA detection with RT-qPCR was also observed in the second mCRPC patient (P12). This indicates an encouraging trend of AR-FL mRNA and ccfDNA presence in patients with high-risk and metastatic disease, matching previous literature Nimir et al. (2019); Stitz et al. (2024). A larger cohort of mCRPC patients would be required to further explore the clinical utility of AR-FL nucleic acid detection at PoC.

**Table 1:**
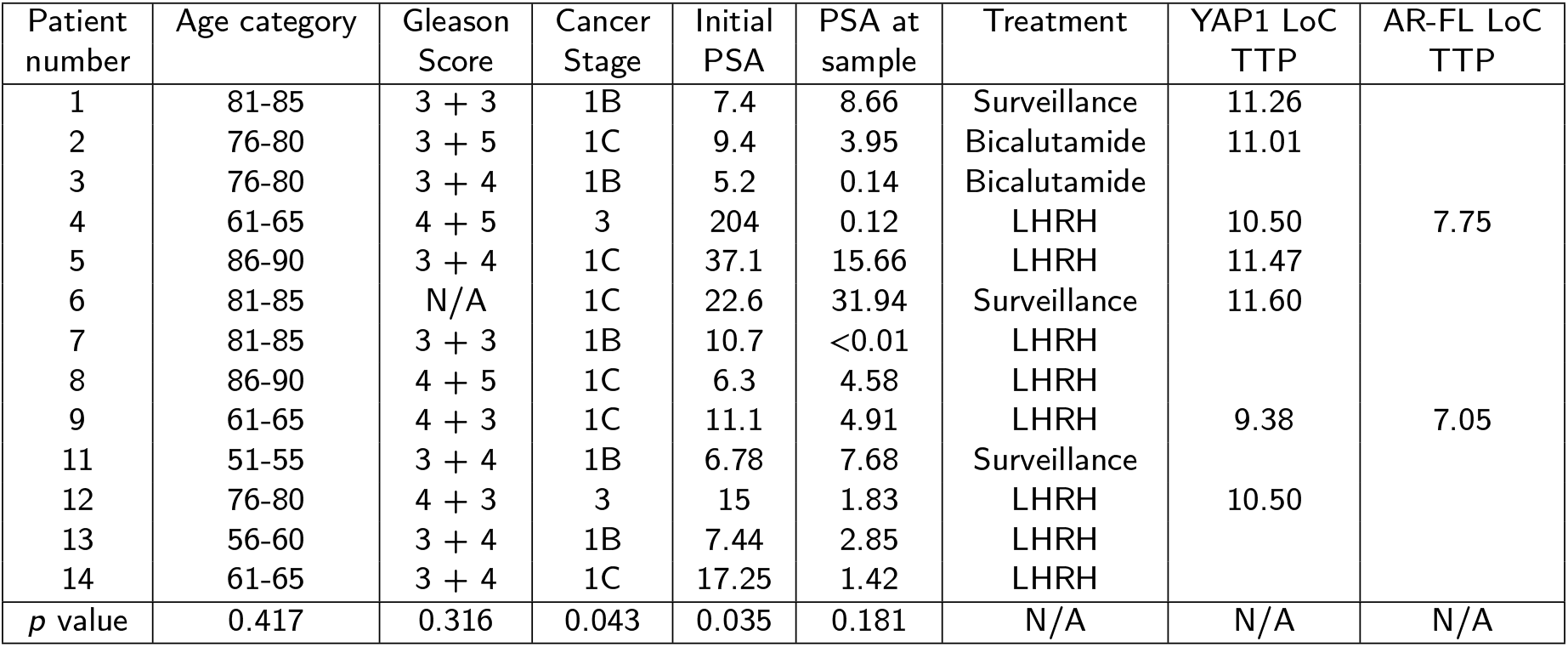
A table of recorded clinical data and the detection of YAP1 nucleic acid and AR-FL mRNA with the ISFET Lab-on-Chip device. The *p* value indicates whether the distinction between YAP1+ and YAP1-cohorts is statistically significant as determined by the Mann-Whitley U test.

| Patient number | Age category | Gleason Score | Cancer Stage | Initial PSA | PSA at sample | Treatment | YAP1 LoC TTP | AR-FL LoC TTP |
| --- | --- | --- | --- | --- | --- | --- | --- | --- |
| 1 | 81-85 | 3 + 3 | 1B | 7.4 | 8.66 | Surveillance | 11.26 |  |
| 2 | 76-80 | 3 + 5 | 1C | 9.4 | 3.95 | Bicalutamide | 11.01 |  |
| 3 | 76-80 | 3 + 4 | 1B | 5.2 | 0.14 | Bicalutamide |  |  |
| 4 | 61-65 | 4 + 5 | 3 | 204 | 0.12 | LHRH | 10.50 | 7.75 |
| 5 | 86-90 | 3 + 4 | 1C | 37.1 | 15.66 | LHRH | 11.47 |  |
| 6 | 81-85 | N/A | 1C | 22.6 | 31.94 | Surveillance | 11.60 |  |
| 7 | 81-85 | 3 + 3 | 1B | 10.7 | <0.01 | LHRH |  |  |
| 8 | 86-90 | 4 + 5 | 1C | 6.3 | 4.58 | LHRH |  |  |
| 9 | 61-65 | 4 + 3 | 1C | 11.1 | 4.91 | LHRH | 9.38 | 7.05 |
| 11 | 51-55 | 3 + 4 | 1B | 6.78 | 7.68 | Surveillance |  |  |
| 12 | 76-80 | 4 + 3 | 3 | 15 | 1.83 | LHRH | 10.50 |  |
| 13 | 56-60 | 3 + 4 | 1B | 7.44 | 2.85 | LHRH |  |  |
| 14 | 61-65 | 3 + 4 | 1C | 17.25 | 1.42 | LHRH |  |  |
| *p* value | 0.417 | 0.316 | 0.043 | 0.035 | 0.181 | N/A | N/A | N/A |

**Figure 3:**
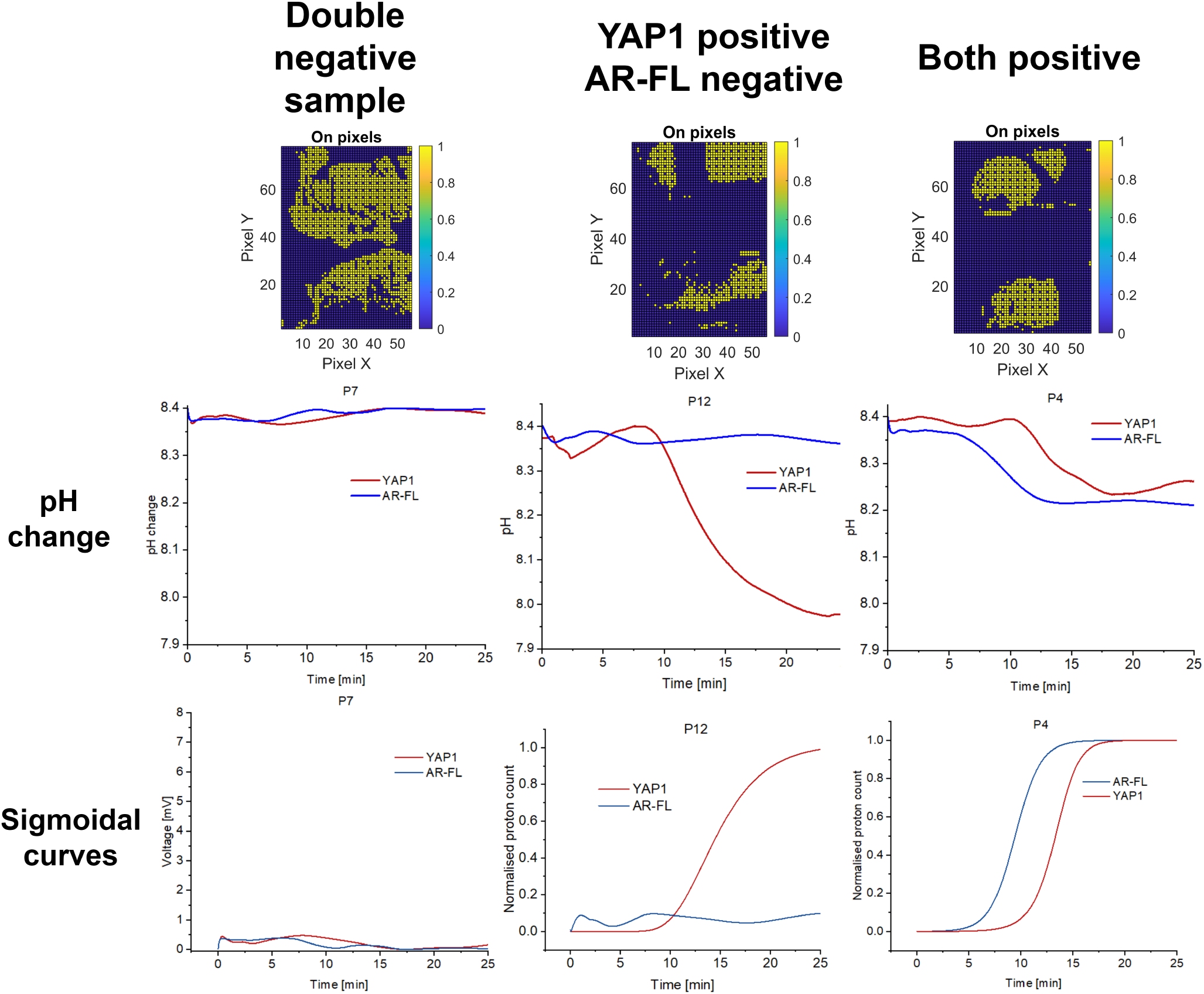
The first frames, pH changes and the sigmoidally-fitted proton count output of the ISFET Lab-on-Chip device for different patient samples. **(a):** P7, where neither YAP1 nucleic acid nor AR-FL mRNA were present as determined by RT-pHLAMP and two step RT-qPCR. **(b)**: P12, where YAP1 nucleic acid was present, but AR-FL mRNA was not present as determined by RT-pHLAMP. **(c):** P4, Where both YAP1 nucleic acid and AR-FL mRNA were present as determined by RT-pHLAMP and two step RT-qPCR reactions.

Circulating AR-FL mRNA detection has been reported previouslyNimir et al. (2019); Lokhandwala et al. (2017). Within the cohort of patients in this work, several patients had metastatic disease (*n = 2*) while the others *(n=11)* were hormone sensitive PCa patients which were considered either intermediate or high risk. A subset of the clinical data is recorded in Table 1. An exhaustive list of clinical data recorded can be found in Supplementary Information Table S2. The clinical staging of PCas was higher in YAP1 positive patients, as determined by RT-pHLAMP assay, compared to YAP1 negative patients (*p* = 0.043, Figure 5 **b**). Patients that have a 1B cancer stage are considered intermediate risk, 1C as high risk and 3 as metastatic. This indicates that the presence of YAP1 nucleic acid, as detected with the LoC device, is associated with metrics for more advanced disease in patient samples. An association with YAP1 nucleic acid presence and initial PSA at time of diagnosis can also be established, where YAP1 nucleic acid presence correlated with higher PSA level at diagnosis (*p* = 0.035, Figure 5 **a**). Our hypothesis anticipated lower concentrations of circulating YAP1 nucleic acid would be present in metastatic patients. However, metastatic sample P4, exhibited some of the fastest TTP values for YAP1 with both RT-qPCR and the ISFET biosensor. Since mCRPC patients have a higher CTC burden than hormone-naive patients, it is likely that the total copy number of YAP1 nucleic acid is higher in these patients, although the total copy number could be lower per cell Nimir et al. (2019); De Bono et al. (2008). Supplementary information Figure S9 further supports this as the relative expression of YAP1 nucleic acid compared to *β*-actin as determined by RT-qPCR ascribes low YAP1 nucleic acid relative expression to the metastatic patients P4 and P12. Further analysis of this in a larger metastatic patient cohort could indicate that YAP1 nucleic acid relative expression could differentiate between high risk and metastatic cancer staging. The introduction of a relevant housekeeping marker for LoC detection in tandem with YAP1 nucleic acid and AR-FL mRNA could additionally provide valuable insight into distinguishing relative copy numbers at PoC.

Once the investigators were unblinded to the clinical data, TRIzol extracted mRNA samples for metastatic patients were additionally tested for AR-V7 with the RT-pHLAMP reaction. Since AR-V7 mRNA is typically a late-stage circulating PCa biomarker, its abundance is low to none in HSPC patient cohorts Nimir et al. (2019); Stitz et al. (2024). As such, the full patient cohort was not tested with the LoC device or benchtop RT-pHLAMP. Instead, the two metastatic patient samples (P4 and P12) and two intermediate risk patients (P11 and P13) were tested with the benchtop AR-V7 RT-pHLAMP reaction (Supplementary Figure S10). P12, while not exhibiting evidence of AR-FL mRNA amplification, was positive for AR-V7 mRNA with both RT-pHLAMP (in 9.66 ± 0.31 min) and RT-qPCR targeting the same mRNA fragment. P4, P11 and P13 showed no evidence of an amplification signal over the 35 min reaction. Simultaneous ISFET LoC detection of AR-FL and AR-V7 mRNA subsequently occurred with P4, P11 and P12 (Figure 4). P11 was included as a negative control, where no AR-FL or AR-V7 mRNA was observed with either RT-pHLAMP or RT-qPCR. Detection of AR-V7 mRNA occurred within 12.55 min with the ISFET LoC device in P12, and no observable deviation from the expected sensor drift occurred with mRNA with TRIzol LS extracted mRNA from P4 and P11 (Figure 4 **c**). AR-FL mRNA in P4 was additionally observed within 8.01 min, compared to 7.75 min when detected simultaneously with YAP1 nucleic acid. Importantly, since the AR-V7 RT-pHLAMP did not render a positive signal in P4, it further indicates the specificity of the reaction to the AR-V7 variant. Shortly after blood sample collection P4 had a radical prostatectomy and P12 had radical external beam radiotherapy. The escalation of treatment for both of these patients aligns with the presence of late-stage markers (AR-FL amplification and AR-V7 mRNA) in their blood samples as detected by the ISFET LoC device.

**Figure 4:**
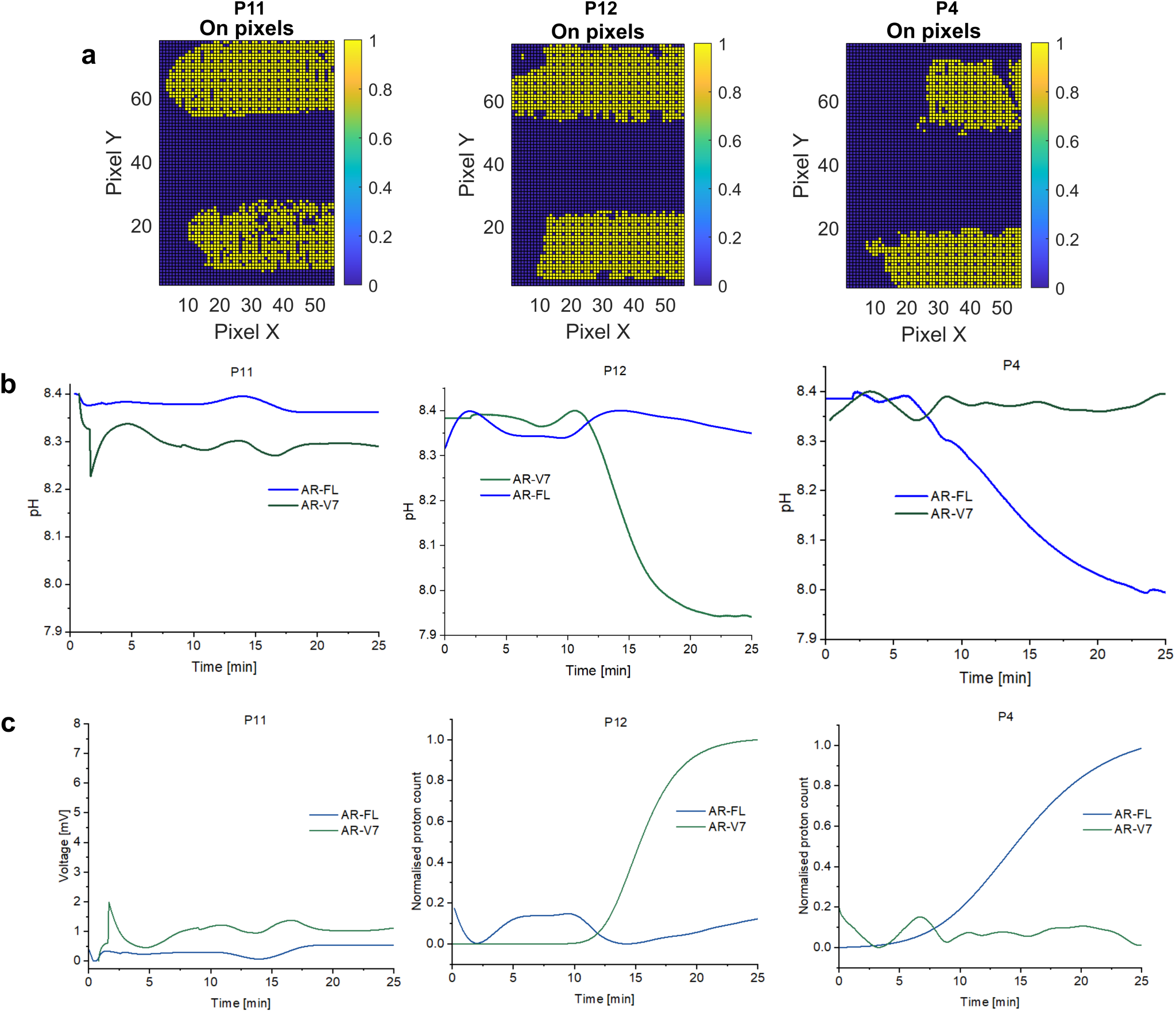
**(a)**: The first frames of the ISFET array surface for dual manifold detection of AR-V7 and AR-FL. **(b)**: The pH changes of ISFET LoC device with the AR-V7 RT-pHLAMP and AR-FL RT-pHLAMP reactions for P11, P12 and P4 TRIzol LS extracted mRNA. **(c)**: The sigmoidal-fitted normalised proton count plots of the AR-V7 and AR-FL sensor outputs.

**Figure 5:**
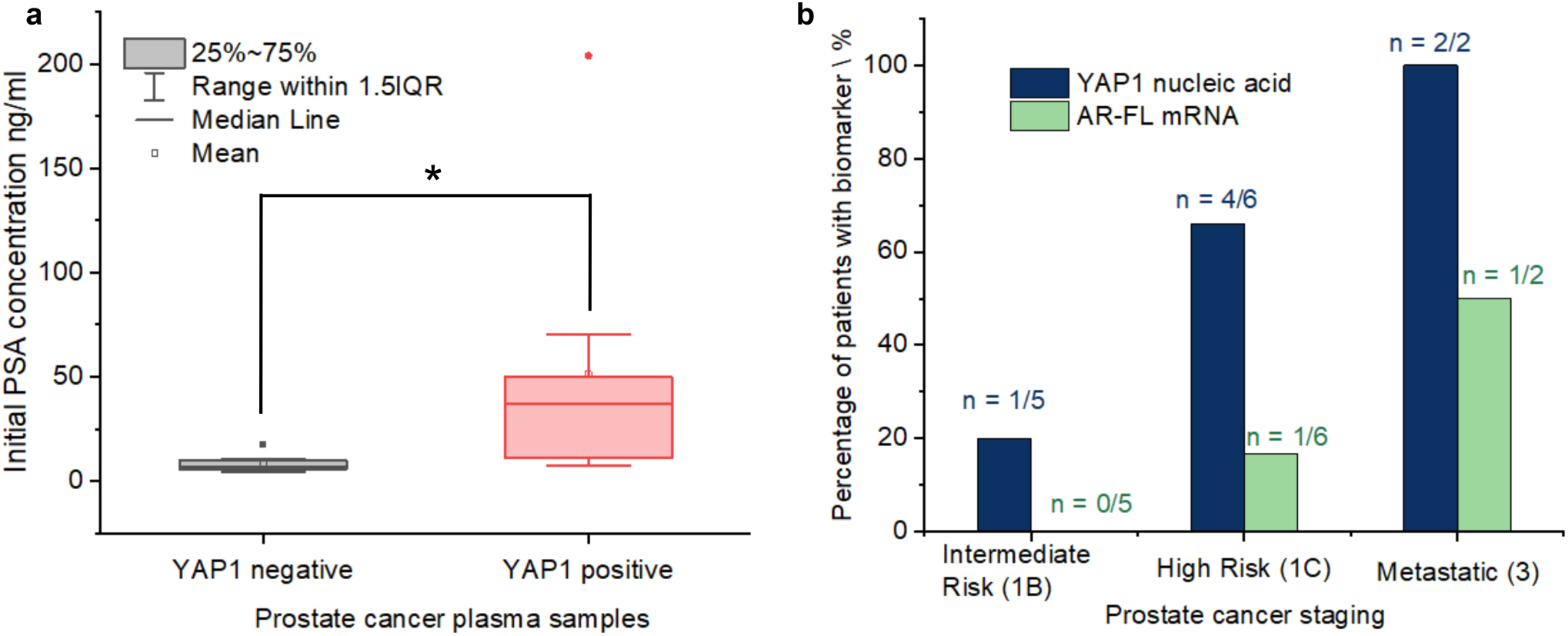
**(a)**: The box plot of YAP1 positive and negative clinical samples as determined by the ISFET LoC device compared to initial PSA levels of the prostate cancer patients at diagnosis. **(b):** YAP1 nucleic acid and AR-FL mRNA presence as determined by the ISFET LoC device in extracted RNA from blood plasma from prostate cancer patients of varying clinical cancer staging.

## Discussion

This work presents the capability for detection of circulating YAP1 nucleic acid and AR-FL and AR-V7 mRNA in the blood of PCa patients using a handheld LoC device for prognostic utility. To the authors’ knowledge, this work is the first ISFET-based biosensor integrated in a Lab-on-Chip, capable of detecting endogenous nucleic acids from the blood of cancer patients in general. It is also the first handheld biosensor capable of detecting circulating mRNA for PCa prognosis. The benchtop reactions and LoC detection of RT-pHLAMP corroborate well, providing further potential for detection of PCa circulating nucleic acids at the point-of-care. Circulating YAP1 nucleic acid presence detected with the LoC device additionally correlated with more advanced cancer staging and initial PSA in a cohort of PCa patients. This indicates that YAP1 nucleic acid ISFET LoC detection can provide clinically actionable information within 15 min after RNA extraction with minimal further processing. YAP1 upregulation has also been observed in multiple solid cancer types including breast, colorectal and liver cancers Guo et al. (2019); Chen et al. (2024); Russell and Camargo (2022). Circulating YAP1 nucleic acid detection at PoC therefore, could provide value to clinicians across several varieties of cancer. AR-FL mRNA amplification and AR-V7 mRNA detection were also achieved with the LoC and these biomarkers were only observed in high risk or metastatic patients.

The chosen extraction methodology for mRNA in this work utilises TRIzol LS reagent. This is currently unsuitable for direct point-of-care extraction. Adaptation of the extraction methodology for PoC compatibility would increase the validity of the device for use within hospitals. Microfluidic approaches could additionally result in a singular device for mRNA extraction from blood plasma followed by detection with RT-pHLAMP and the ISFET biosensor.

Further detection of AR-V7 mRNA, AR-FL mRNA amplification and YAP1 nucleic acid within a larger cohort of patients would provide more information on their prognostic value. Previous work has additionally developed detection of TMPRSS2-ERG mRNA with a RT-pHLAMP assay Broomfield et al. (2023). Simultaneous detection of a panel of four markers (TMPRSS2-ERG, AR-FL, AR-V7 and YAP1) would provide further information on tumour state and counter heterogeneity between PCa patients. At PoC, detection of these markers could rapidly provide information to clinicians and result in changes to medication or treatment. Further exploration of YAP1 nucleic acid expression relative to a housekeeping gene could also allow for greater differentiation of high risk and metastatic patients. Implementation of a housekeeping RT-pHLAMP reaction onto the LoC device could aid with relative expression of YAP1 presence at PoC. This work increases the potential for personalised PCa prognostics to occur rapidly, reliably and sensitively within the clinic by non-specialised personnel.

## 3. Experimental

### RT-pHLAMP

Primer sequences and methods have previously been reported for YAP1 RT-pHLAMP. Bovine Serum Albumin (NEB, B9000S) was discontinued in 2022 and was replaced with Recombinant Albumin (NEB, B9200S) for all RT-pHLAMP experiments conducted after this date.

The updated protocol for AR-FL RT-pHLAMP reactions is recorded here. All reactions were completed in quadruplicate. Each 10 *μ*L experiment contained: 1 *μ*L of customized isothermal buffer, 0.25 *μ*L of MgSO4 (100 mM stock), 1.4 *μ*L of dNTPs (10 mM stock of each nucleotide), 0.9 *μ*L of BSA (20 mg/mL stock), 0.25 *μ*L of SYTO 9 green (20 *μ*M stock), 0.125 *μ*L of NaOH (0.2 M stock), 0.042 *μ*L of Bst 2.0 WarmStart DNA polymerase (120,000 U/mL stock, NEB), 0.3 *μ*L of WarmStart RTx reverse transcriptase (15,000 U/mL stock, NEB), 1 *μ*L of 10× LAMP primer mix (30 *μ*M FIP and BIP, 15 *μ*M LB and LF, 3.75 *μ*M F3 and B3), 1 *μ*L of RNA sample, and the remaining solution was topped up to 10 *μ*L with nuclease-free water.

All benchtop RT-pHLAMP experiments were conducted with a StepOnePlus Real-Time PCR machine (Thermofisher) and run for 70 cycles of 30 s at 63 ^0^C. Determination of TTP values was established with a post-processing algorithm which normalised amplification curves and defined TTP as the time taken to cross a normalised fluorescent value of 0.2. This is in accordance with standard recording of fluorescent curves from thermal cycling equipment.

### Blood collection and storage

Blood samples were extracted using either 4 mL or 6 mL EDTA vacutainer tubes by nurse practitioners from Charing Cross Hospital, London. Blood samples were immediately placed on ice. Within 4 hours of blood collection, the samples were centrifuged at 1200 *g* for 10 min at 4 °C. The plasma supernatant was stored at −70 °C in preparation for downstream applications.

### TRIzol RNA Extraction

RNA extraction was conducted with TRIzol™ LS Reagent (Thermofisher) according to the manufacturer’s protocol with the following adjustments. 250 *μ*L of plasma was utilised for each extraction. Isoproponol and ethanol were cooled in dry ice in preparation for the respective precipitation and washing steps. Resuspension of the RNA pellet occurred with 25 *μ*L of nuclease-free water. The resultant solution was not incubated at 55-60 °C and was instead immediately stored at −70 °C in preparation for RNA experiments. All TRIzol RNA extractions were run with a nuclease-free water control. Pooled mixed-male citrated plasma (PR200M, TCS Biosciences) was utilised for synthetic RNA spiked extractions.

### cDNA formation and qPCR

RNA to cDNA conversion was conducted using a RevertAid First Strand cDNA synthesis Kit (ThermoFisher) using 4 *μ*L of extracted RNA and random primers. The optional GC rich region incubation at 65 °C for 5 min was conducted. cDNA was either utilised immediately for qPCR or stored at −70 °C.

qPCR reactions were conducted with a StepOnePlus thermal cycler (Thermofisher). All experiments were run for 50 cycles and technical repeats were completed in quadruplicate. For qPCR positive controls, 4 *μ*L of 1 ng / *μ*L LNCaP cell line mRNA was converted to cDNA. The following experimental conditions were conducted for *β*-actin and YAP1 primer set 2. The annealing temperature for the AR-FL small fragment reactions was 65 °C but otherwise the same as the list below. For these assays, forward and back primers were present at a concentration of 250 nM per reaction.

- 95 °C for 2 minutes
- 60 °C for 30 seconds
- 95 °C for 1 second
- Steps 1. and 2. were repeated for 50 cycles
- Melting Curve: 60 seconds at 60 °C followed by data recording every 0.3 °C until 95°C.

For YAP1 qPCR primer set 1, AR-V7 qPCR and AR-FL large fragment assays the annealing time was adjusted to 1 minute per cycle. qPCR primer concentrations for the YAP1 primer set 1, AR-V7 and AR-FL large fragment assays were 600 nM, 600 nM and 1000 nM respectively. Annealing temperatures of these assays was 57 °C, 60 °C and 60 °C respectively. Optimisation of annealing temperature and primer concentrations was established experimentally.

Analysis of Cq values from all qPCR reactions was determined with a MATLAB algorithm. Normalisation of amplification curves occurred and determination of Cq values occurred when the normalised fluorescence threshold of 0.1 was reached. This threshold fluorescence value was utilised as positive and negative samples could be comfortably distinguished at this fluorescence threshold. Quadruplicate averages that were recorded above 40 cycles were determined as negative. For the AR-V7 RT-qPCR reaction, this was lowered to 38 cycles due to the presence of off-target amplification at late cycle numbers.

Each 10 *μ*L qPCR reaction contained 5 *μ*L of FAST SYBR Green Master Mix (Applied Biosystems), 2 *μ*L of cDNA sample, 0.5 *μ*L of reverse primer (x20 stock), 0.5 *μ*L of forward primer (x20 stock), 2 *μ*L Nuclease-free water.

A comprehensive list of primers utilised for both qPCR and LAMP can be found in the Supplementary information section Table S1.

### Lab-on-Chip detection

Single-well LoC testing was conducted identically to the procedure provided here Broomfield et al. (2022a). For dual-well testing with the ISFET biosensor a purpose-built 5 mm acrylic manifold was utilised. Reaction chamber wells were reduced to 15 *μ*L. The passivation layer of the ISFET pixels was adjusted to a higher *κ* dielectric material. A dual-hole adhesive gasket (Double-sided smooth lamination filmic tape, Tesa®) and bolting of the manifold to the PCB reduced the likelihood of assay solution leakage. A single 0.01 mm Ag/AgCl chlorodised reference electrode was threaded through both wells and soldered to a contact point. Nuclease-free water was pipetted in the reaction chamber for 700s prior to the reaction start. The top well nuclease-free water was replaced first with AR-FL RT-pHLAMP reaction and sample. Secondly, the bottom well was replaced with either AR-V7 or YAP1 RT-pHLAMP reaction. Once the sensor output had stabilised, data was recorded. Since the injection of the RT-pHLAMP reactions occurred at different time-points the beginning of the reactions differed by around 30 s. All dual-well reactions were run for 25 min to ensure all amplification had terminated. For P9, sensor output recording was halted at 20 min. However, on account of the fast TTP values of this sample, all amplification had subsequently occurred. pH change (Sentron S1600 pH meter and MicroFET pH probe) and DNA quantification (Qubit 3.0 fluorometer, Invitrogen) post-reaction were recorded and utilised to confirm positive and negative reactions.

### 3.1 Statistical Analyses

The unpaired two-tailed Student’s t-test was used to determine the statistical significance of differences in YAP1 RT-pHLAMP TTPs between clinical samples. The Mann-Whitley U non-parametric test was utilised for all subsequent statistical analyses of clinical data. This statistical test was used since clinical staging and Gleason Scoring for example are ordinal, where the t-test cannot be used. In the cases where the data was quantitative (IPSA for example) it was assumed that the clinical data would not be a standard distribution and therefore utilisation of the parametric t-test was excluded. A *p* < 0.05 was defined as the boundary for statistical significance.

### 3.2 Ethical approval

Informed consent for blood samples from prostate cancer patients was taken in accordance with NIHR guidelines. Ethical approval for blood sample extraction was sought and ascertained from the Imperial College NHS Health-care Trust. Blood samples were taken alongside standard PSA blood testing. Non-clinical co-authors were blinded to patient data and severity of disease. Patient data was anonymised and maintained under the NHS trust database. The clinical samples were recorded under the sub-collection URO CB 13 029 with the Imperial College Healthcare NHS Trust Tissue Bank.

## Supporting information

Supplementary information: Further experimental and clinical data

## Data Availability

All data produced in the present study are available upon reasonable request to the authors.

## 3.3 Acknowledgements

The authors thank all members of the Georgiou and Bevan laboratories for insightful discussions. This work was funded in part by a Convergence Science PhD studentship (C24523) and in part by Prostate Cancer UK grants MA-COE18-001 and Grant RIA17-ST2-017. Infrastructure support was provided by Imperial Experimental Cancer Medicine Centre, National Institute for Health Research (NIHR) Imperial Biomedical Research Centre (BRC) and Imperial College Healthcare NHS Trust Tissue Bank.

## 3.4. Data Statement

Research data contains sensitive and/or confidential patient data and therefore has not been made available in a repository.

## CRediT authorship contribution statement

**Joseph Broomfield:** Conceptualisation, Investigation, Clinical data analysis and writing-Original draft. **Melpomeni Kalofonou:** Conceptualisation, supervision, writing-review and editing, funding acquisition.. **Costanza Gulli:** Investigation, writing-review and editing. **Sue M. Powell:** Investigation, writing-review and editing. **Rayzel C. Fernandes:** Investigation, writing-review and editing. **Damien A.Leach:** Clinical data analysis, writing-review and editing. **Nicolas Moser:** Resources, methodology, writing-review and editing. **Naveed Sarwar:** Clinical sample acquisition, Writing-review and editing. **Stephen Mangar:** Conceptualisation, Clinical sample acquisition, clinical data analysis, writing-review and editing. **Charlotte L. Bevan:** Conceptualisation, Funding acquisition, methodology, supervision, writing-review and editing. **Pantelis Georgiou:** Conceptualisation, Funding acquisition, methodology, supervision, writing-review and editing.

