## Supplementary information: Further experimental and clinical data for "Handheld ISFET Lab-on-Chip detection of YAP1 nucleic acid and AR-FL and AR-V7 mRNA from Liquid Biopsies for Prostate Cancer Prognosis"

#### Contents

**1 YAP1 and AR-FL RT-pHLAMP detection from synthetic RNA spiked into commercial plasma. 2**

**2 YAP1 RT-qPCR primer set 1 optimisation 3**

**3 AR-FL RT-pHLAMP assay optimisation 4**

**4 AR-FL Large fragment qPCR optimisation 5**

**5 Single-well manifold testing with clinical samples and the YAP1 RT-pHLAMP assay and clinical data 5**

**6 RT-qPCR and RT-pHLAMP primer sequences 8**

**7 Lab-on-Chip detection of all clinical samples 9**

**8 YAP1 RT-pHLAMP correlation between LoC and benchtop TTPs 11**

**9 YAP1 expression relative to beta actin and YAP1 RT-qPCR primer set 1 12**

**10 AR-V7 mRNA RT-qPCR and RT-pHLAMP benchtop detection and PSA levels. 13**

### 1 YAP1 and AR-FL RT-pHLAMP detection from synthetic RNA spiked into commercial plasma.

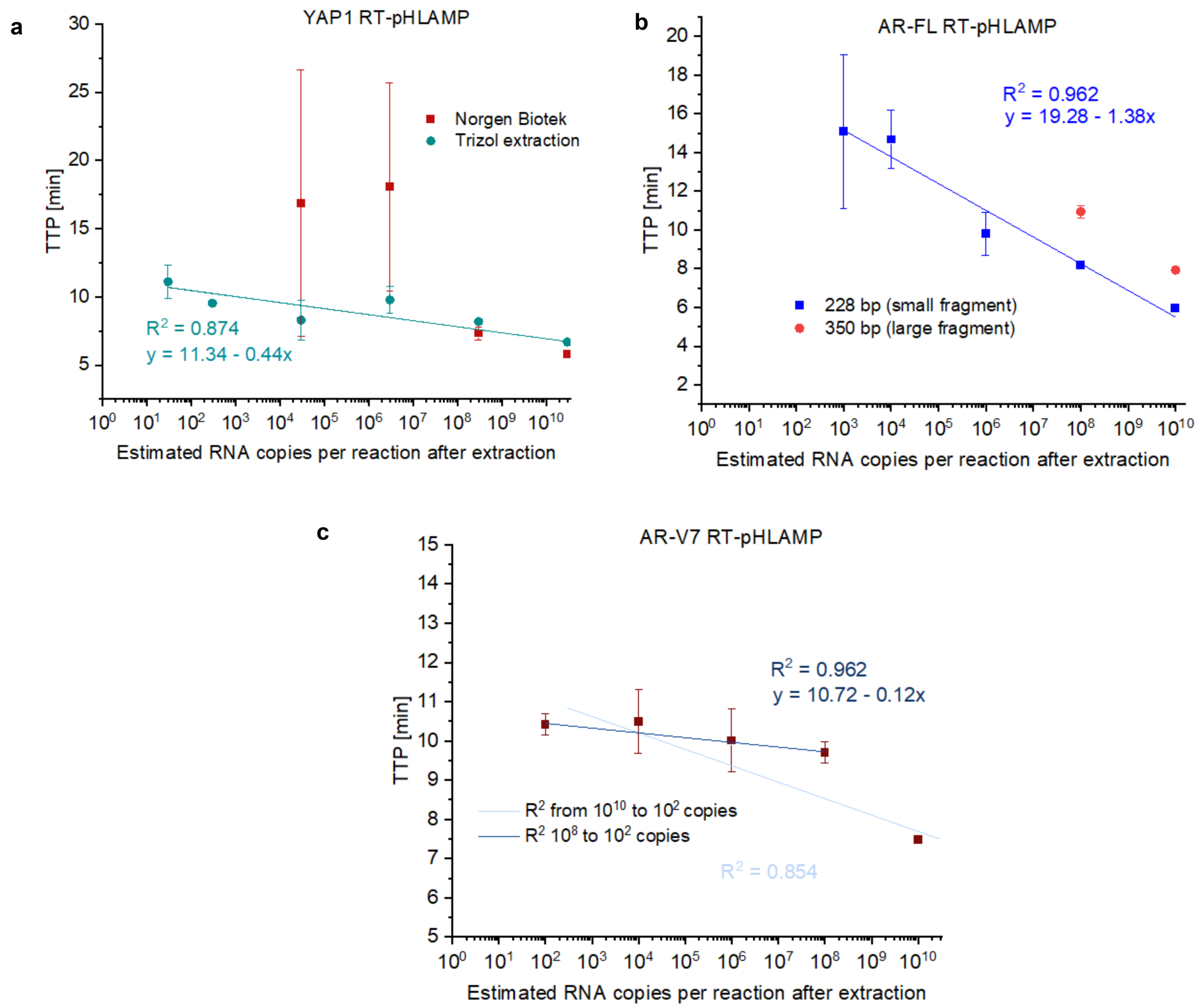

Figure S1: **(a):** The comparison of extraction methodologies for RNA from blood with the YAP1 RT-pHLAMP reaction. Synthetic YAP1 RNA was spiked into commercial mixed male plasma and extracted with either TRIzol LS reagent with the conditions described in the main paper experimental section or the Norgen Biotek plasma/serum RNA purification mini kit. **(b):** The AR-FL RT-pHLAMP reaction detection of small (228 bp) and large (350bp) synthetic RNA fragments when spiked into mixed male plasma and extracted with TRIzol LS reagent. **(c):** The AR-V7 RT-pHLAMP reaction detection of synthetic AR-V7 RNA when spiked into mixed male plasma and extracted with TRIzol LS reagent.

#### 2 YAP1 RT-qPCR primer set 1 optimisation

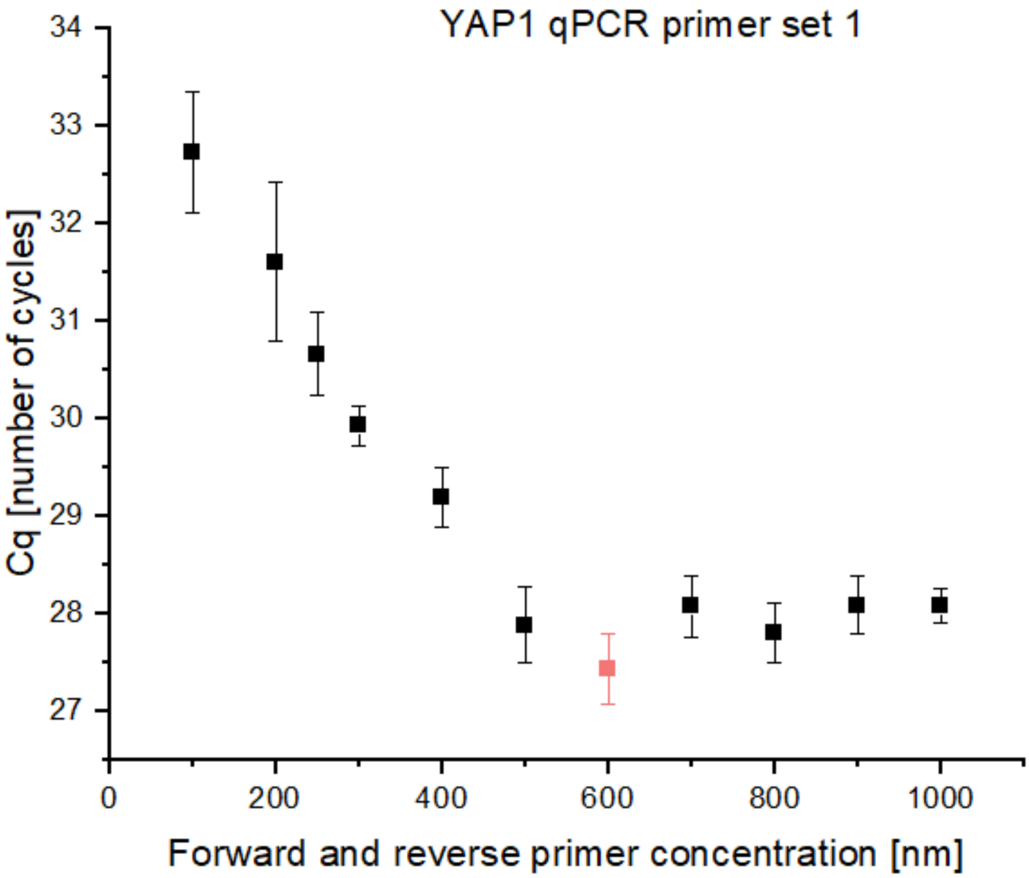

Figure S2: Optimisation of the YAP1 RT-qPCR primer set 1. Different concentrations of forward and reverse primers were utilised. 600 nm provided the fastest Cq value and so was utilised for all future experiments.

### 3 AR-FL RT-pHLAMP assay optimisation

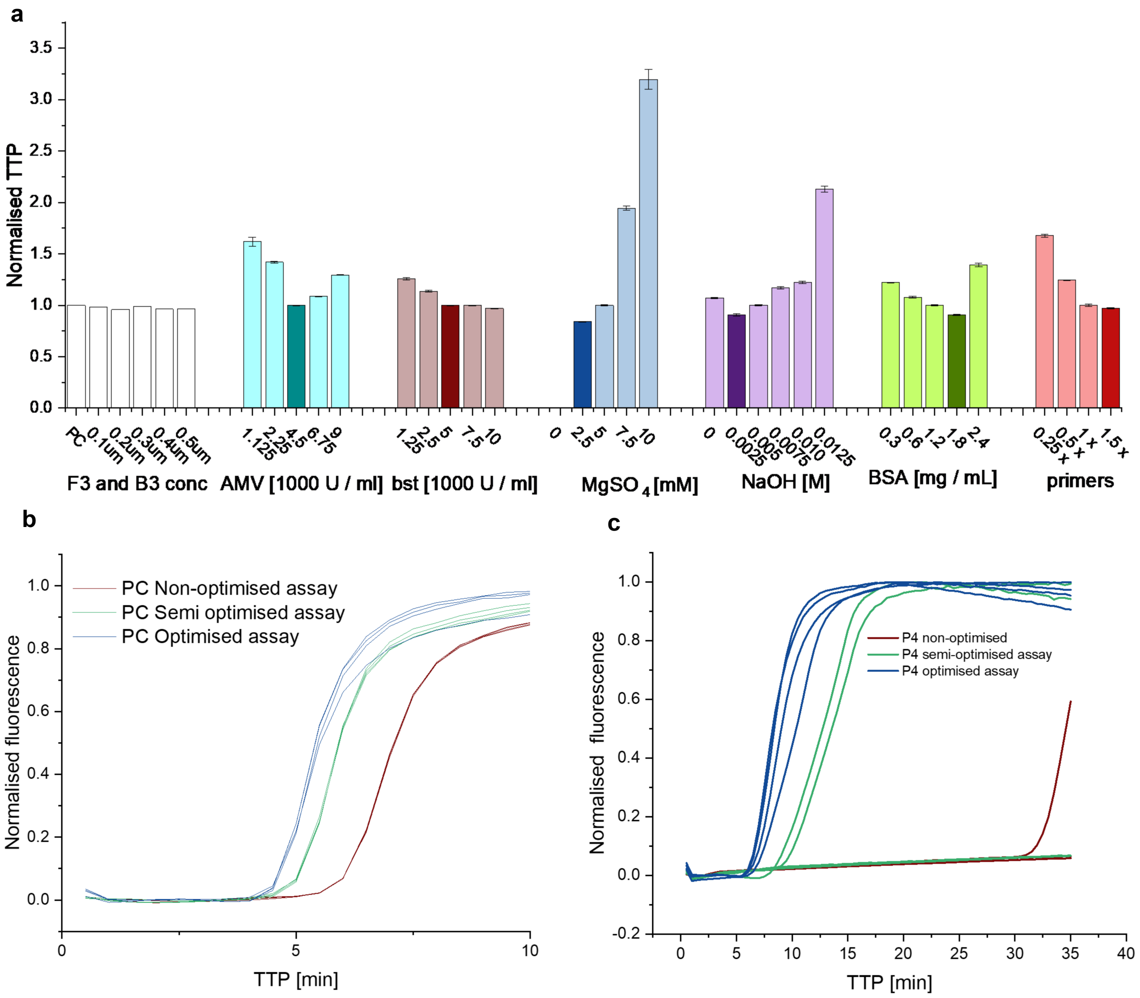

Figure S3: **(a)**: Optimisation of the AR-FL RT-pHLAMP assay. Colours shown in bold colours were reaction conditions that were taken forward for clinical sample testing. The fastest TTP concentration was taken forward in most cases. **(b)**: The normalised fluorescence amplification curve of the pre-optimised, semi-optimised and fully-optimised AR-FL RT-pHLAMP assay. The semi-optimised reaction only had NaOH and $MgSO_4$ concentrations adjusted from the assay conditions published here [1]. **(c)**: The normalised fluorescence curves of the AR-FL RT-pHLAMP assay on extracted RNA from the blood plasma of P4. Quadruplicate detection within 10 min was realised in the fully optimised assay.

### 4 AR-FL Large fragment qPCR optimisation

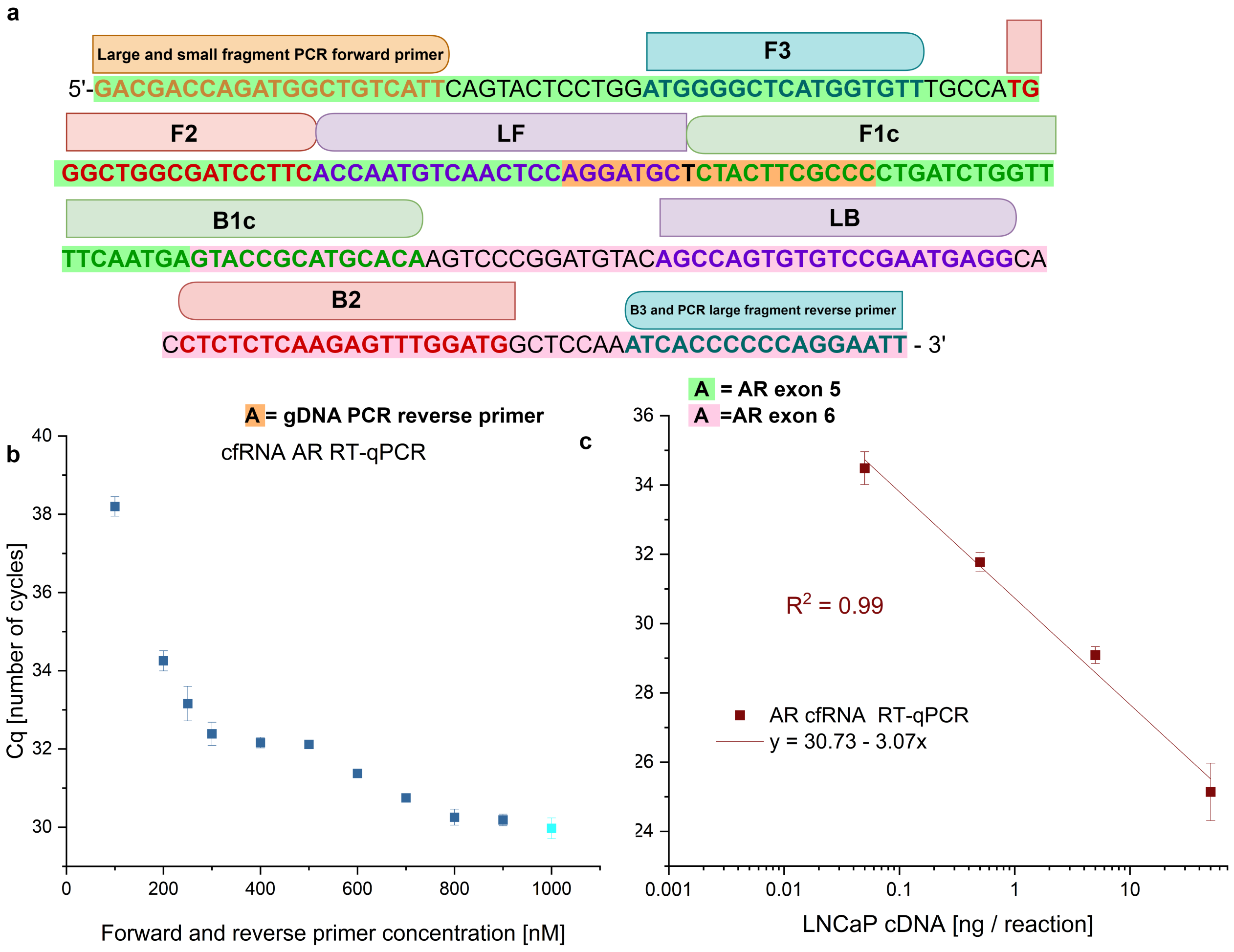

Figure S4: **(a)**: The region of AR-FL DNA that is detected by the RT-pHLAMP and the small and large fragment AR-FL RT-qPCR assays. **(b)**: The primer concentration optimisation of the large fragment AR-FL RT-qPCR assay. The fastest cycle detection of 0.5 ng of LNCaP RNA was with forward and reverse primer concentrations of 1000 nM per reaction. **(c)**: The sensitivity and quantitative nature of the optimised large fragment AR-FL RT-qPCR assay. Detection of AR-FL mRNA was observed down to 0.05 ng per reaction with a highly quantitative assay.

### 5 Single-well manifold testing with clinical samples and the YAP1 RT-pHLAMP assay and clinical data

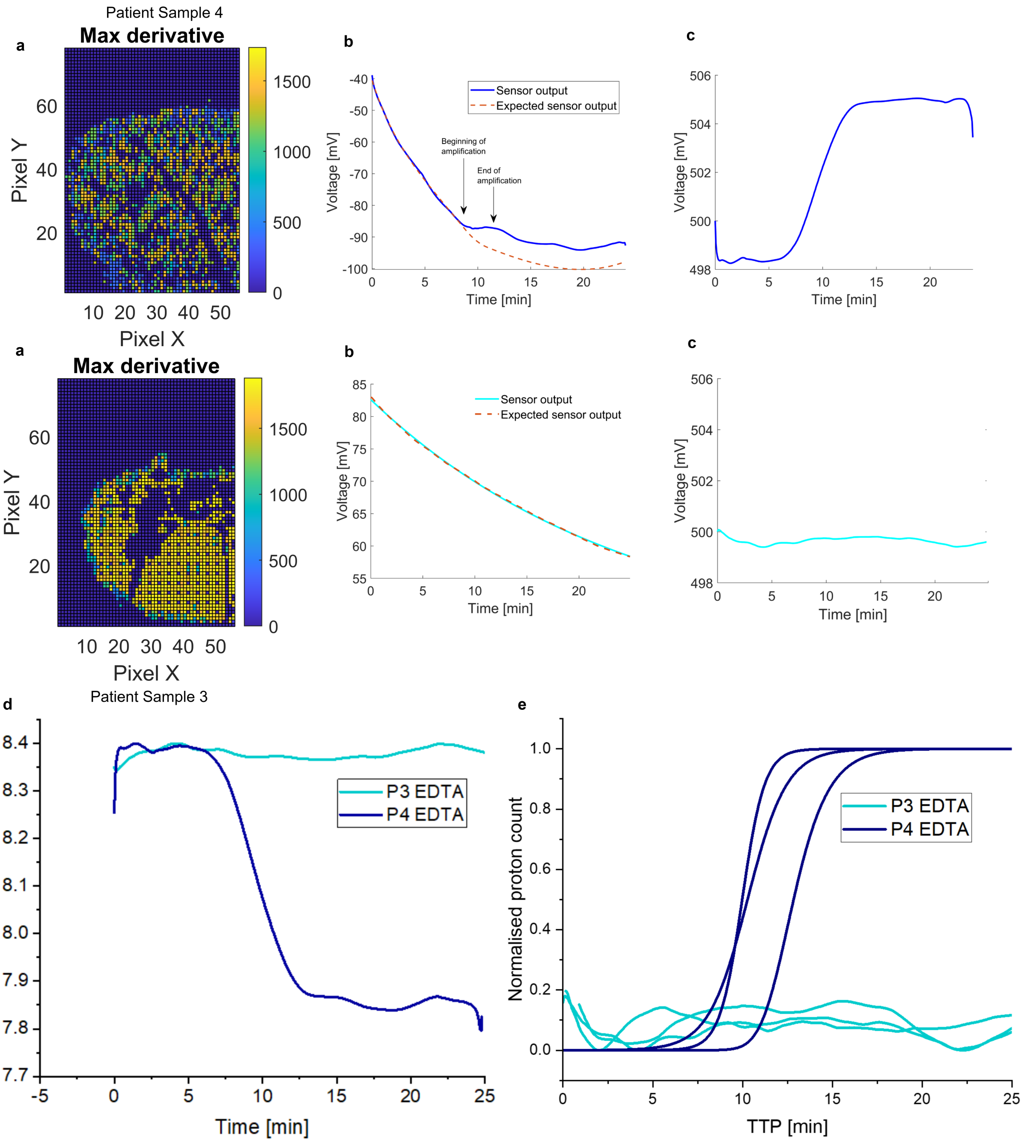

Figure S5: Detection of positive and negative YAP1 nucleic acid samples using the LoC device. **(a)** The first frame of the ISFET array indicating which pixels were in range to detect pH change. **(b)** The sensor outputs of P3 and P4 which had been determined as negative and positive for YAP1 nucleic acid respectively by RT-qPCR and RT-pHLAMP. **(c)**: The change in voltage between expected sensor output and the actual sensor output of the ISFET biosensor. **(d)**: The pH change detected by the ISFET biosensor for a P3 and P4 patient sample. **(e)**: The sigmoidally-fitted normalised proton count output of the biosensor for triplicate P3 and P4 patient samples.

| Patient number | trial number | age category at date of sample | T | N | M | GS | GS | IPSA | DATE OF SAMPLE |
|---|---|---|---|---|---|---|---|---|---|
| 1 | P1 | 81-85 | 1 | 0 | 0 | 3 | 3 | 7.4 | 10.02.2023 |
| 2 | P2 | 76-80 | 3 | 0 | 0 | 3 | 5 | 9.39 | 10.02.2023 |
| 3 | P3 | 76-80 | 2 | 0 | 0 | 3 | 4 | 5.2 | 10.02.2023 |
| 4 | P4 | 61-65 | 3 | 1 | 1 | 4 | 5 | 204 | 10.02.2023 |
| 5 | P5 | 86-90 | 2 | 0 | 0 | 3 | 4 | 37.12 | 24.02.2023 |
| 6 | P6 | 81-85 | 3 | 0 | 0 | x | x | 22.55 | 24.02.2023 |
| 7 | P7 | 81-85 | 2 | 0 | 0 | 3 | 3 | 10.7 | 24.02.2023 |
| 8 | P8 | 86-90 | 3 | 0 | 0 | 4 | 5 | 6.3 | 24.02.2023 |
| 9 | P9 | 61-65 | 3 | 0 | 0 | 4 | 3 | 11.09 | 24.02.2023 |
| | | | | | | | | | -- |
| 11 | P11 | 51-55 | 3 | 0 | 0 | 3 | 4 | 6.78 | 27.02.2023 |
| 12 | P12 | 76-80 | 3 | 1 | 1 | 4 | 3 | 15 | 27.02.2023 |
| 13 | P13 | 56-60 | 2 | 0 | 0 | 3 | 4 | 7.44 | 27.02.2023 |
| 14 | P14 | 61-65 | 3 | 0 | 0 | 3 | 4 | 17.25 | 27.02.2023 |

| Patient number | Trial Number | PSA AT TIME OF SAMPLE | STAGE AT TIME OF SAMPLE 1A LOW RISK, 1B INTERMEDIATE RISK 1C HIGH RISK, 3 METASTATIC | TREATMENT AT TIME OF SAMPLE | DURATION OF TREATMENT AT TIME OF SAMPLE (MONTHS) | DIABETES 1-N0 2- YES |
|---|---|---|---|---|---|---|
| 1 | P1 | 8.66 | 1B | SURVEILLANCE | 0 | 1 |
| 2 | P2 | 3.95 | 1C | BICALUTAMIDE | 3 | 2 |
| 3 | P3 | 0.14 | 1B | BICALUTAMIDE | 12 | 1 |
| 4 | P4 | 0.12 | 3 | LHRH | 8 | 2 |
| 5 | P5 | 15.66 | 1C | LHRH | 7 | 1 |
| 6 | P6 | 31.94 | 1C | SURVEILLANCE | 0 | 1 |
| 7 | P7 | <0.01 | 1B | LHRH | 55 | 1 |
| 8 | P8 | 4.58 | 1C | LHRH | 1 | 2 |
| 9 | P9 | 4.91 | 1C | LHRH | 0 | 2 |
| 11 | P11 | 7.68 | 1B | SURVEILLANCE | 0 | 1 |
| 12 | P12 | 1.83 | 3 | LHRH | 19 | 2 |
| 13 | P13 | 2.85 | 1B | LHRH | 14 | 1 |
| 14 | P14 | 1.42 | 1C | LHRH | 20 | 1 |

| Patient number | Trial Number | COPD 1-NO 2-YES | IHD/AF/PAC 1-N0 2-YES | HYPERTENSION 1-NO 2-YES | SMOKER 1-NO 2 YES | EX SMOKER | PREVIOUS PROSTATE RT 1-NO 2-YES |
|---|---|---|---|---|---|---|---|
| 1 | P1 | 2 | 1 | 2 | 1 | | 1 |
| 2 | P2 | 1 | 2 | 2 | 1 | | 1 |
| 3 | P3 | 1 | 1 | 1 | 1 | | 1 |
| 4 | P4 | 1 | 1 | 1 | 1 | | 1 |
| 5 | P5 | 1 | 1 | 2 | 1 | | 1 |
| 6 | P6 | 1 | 2 | 1 | 1 | | 1 |
| 7 | P7 | 2 | 1 | 1 | 1 | | 2 |
| 8 | P8 | 1 | 2 | 2 | 1 | EX | 1 |
| 9 | P9 | 1 | 1 | 2 | 1 | EX | 1 |
| 11 | P11 | 1 | 1 | 1 | 1 | EX | 1 |
| 12 | P12 | 1 | 1 | 2 | 1 | EX | 2 |
| 13 | P13 | 1 | 1 | 2 | 1 | | 1 |
| 14 | P14 | 2 | 1 | 1 | 2 | | 2 |

Table S1: Clinical data of patients P1-P14. Patient P10 was not utilised and so has been omitted from analysis. The following abbreviations are used in the table. T, N and M come from TNM staging. GS = Gleason Score. IPSA = initial PSA at diagnosis. COPD = chronic obstructive pulmonary disease. IHD = ischaemic heart disease. AF = atrial fibrillation. PAC = premature atrial contraction. RT = Radiotherapy.

### 6 RT-qPCR and RT-pHLAMP primer sequences

| Primers | Primer sequence |
|---|---|
| **AR-FL RT-pHLAMP** | |
| F3 | ATGGGGCTCATGGTGTT |
| B3 | AATTCCTGGGGGGTGATT |
| FIP | AACCAGATCAGGGGCTGGCGATCCTTCACC |
| BIP | TTCAATGAGTACCGCATGCACAAGTGC |
| | CATCCAAACTCTTGAGAGAGGTG |
| LF | GCATCCTGGAGTTGACATTGGT |
| LB | AGCCAGTGTGTCCGAATGAGG |
| **YAP1 RT-pHLAMP** | |
| F3 | TTTGCCCAGTTATACCTCA |
| B3 | CAAGAAGCAGTTAAGCACTT |
| FIP | TCAGTACAGAGGGCATCGTTAGCAGTACTGTGATACCT |
| BIP | CCTGAAGGAGACCTAAGAGTCAGGACATAAAACAAGAGACCA |
| LF | CAAAGCACTGTGCCAGGT |
| LB | CCCTTTTTGAGTTTGAATCATAGCC |
| **AR-V7 RT-pHLAMP** | |
| F3 | CTAGCCTTCTGGATCCCA |
| B3 | AGGCTAGATGTAAGAGGGA |
| FIP | TTCTGTGGATCAGCTACTAACCTAGATCTTAGCCTCAG |
| BIP | AGTAAACAAGGACCAGATTTCTGTAGTCTCTCAGTGTGTTTGA |
| LF | GCTCAGTGACAGGGCCTGAG |
| LB | CCAGGAGAAGAAGCCAGCCA |
| **YAP1 RT-qPCR primer set 1** | |
| Forward | TTTGCCCAGTTATACCTCA |
| Reverse | CAAGAAGCAGTTAAGCACTT |
| **YAP1 RT-qPCR primer set 2** | |
| Forward | GCACCTCTGTGTTTTAAGGGTCT |
| Reverse | CAACTTTTGCCCTCCTCCAA |
| **AR-FL RT-qPCR small fragment** | |
| Forward | GACGACCAGATGGCTGTCATT |
| Reverse | GGGCGAAGTAGAGCATCCT |
| **AR-FL RT-qPCR large fragment** | |
| Forward | GACGACCAGATGGCTGTCATT |
| Reverse | AATTCCTGGGGGGTGATT |
| **AR-V7 RT-qPCR large fragment** | |
| Forward | CTAGCCTTCTGGATCCCA |
| Reverse | AGGCTAGATGTAAGAGGGA |
| **$\beta$- actin RT-qPCR primers** | |
| Forward | GGCATCCTCACCCTGAAGTA |
| Reverse | GGTCATCTTCTCGCGGTTG |

Table S2: Primer sequences for RT-qPCR and RT-pHLAMP reactions. Sequences are shown 5' to 3'.

### 7 Lab-on-Chip detection of all clinical samples

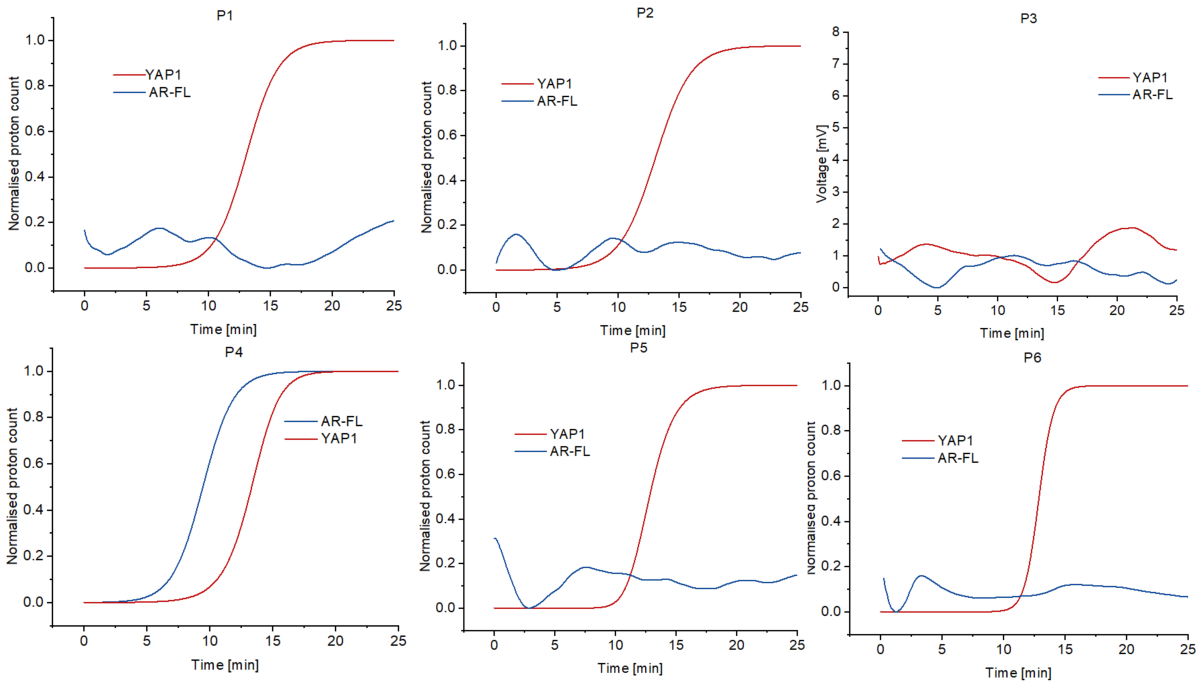

Figure S6: P1-P6 clinical samples on the LoC device. Samples with positives in YAP1 or both YAP1 and AR-FL are normalised. Double negatives remain as the voltage output of the expected sensor drift subtracted from the sensor output.

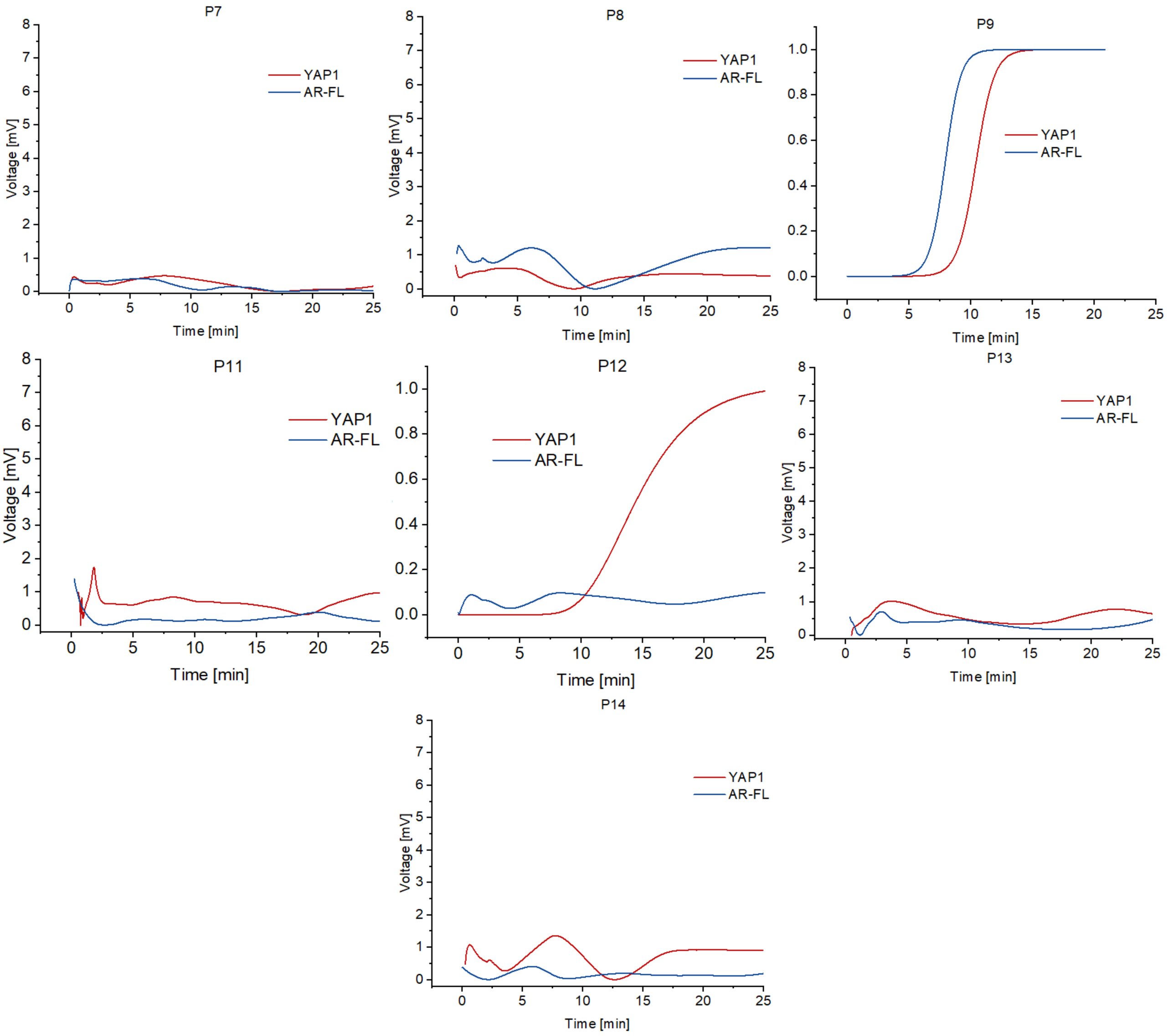

Figure S7: P7-P14 clinical samples with positives in YAP1 or both YAP1 and AR-FL are normalised. Double negatives remain as the voltage output of the expected sensor drift subtracted from the sensor output.

#### 8 YAP1 RT-pHLAMP correlation between LoC and benchtop TTPs

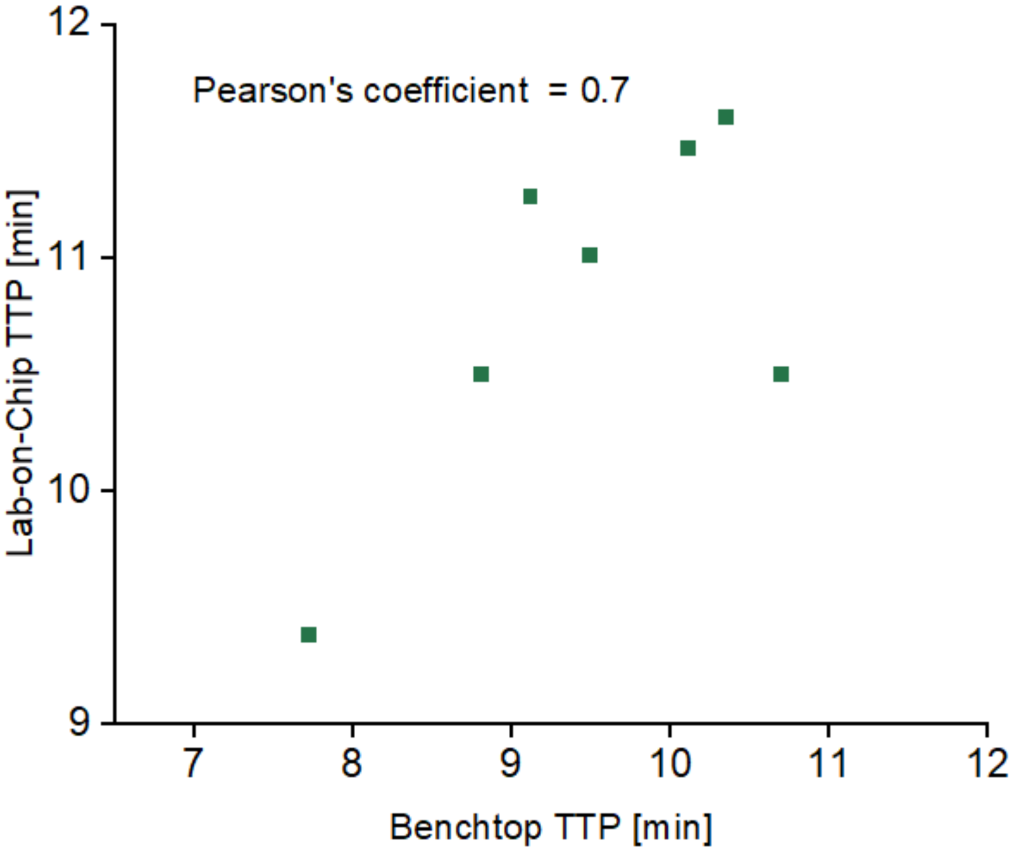

Figure S8: The correlation of benchtop to Lab-on-Chip TTP values on clinical samples with the YAP1 RT-pHLAMP assay.

#### 9 YAP1 expression relative to beta actin and YAP1 RT-qPCR primer set 1

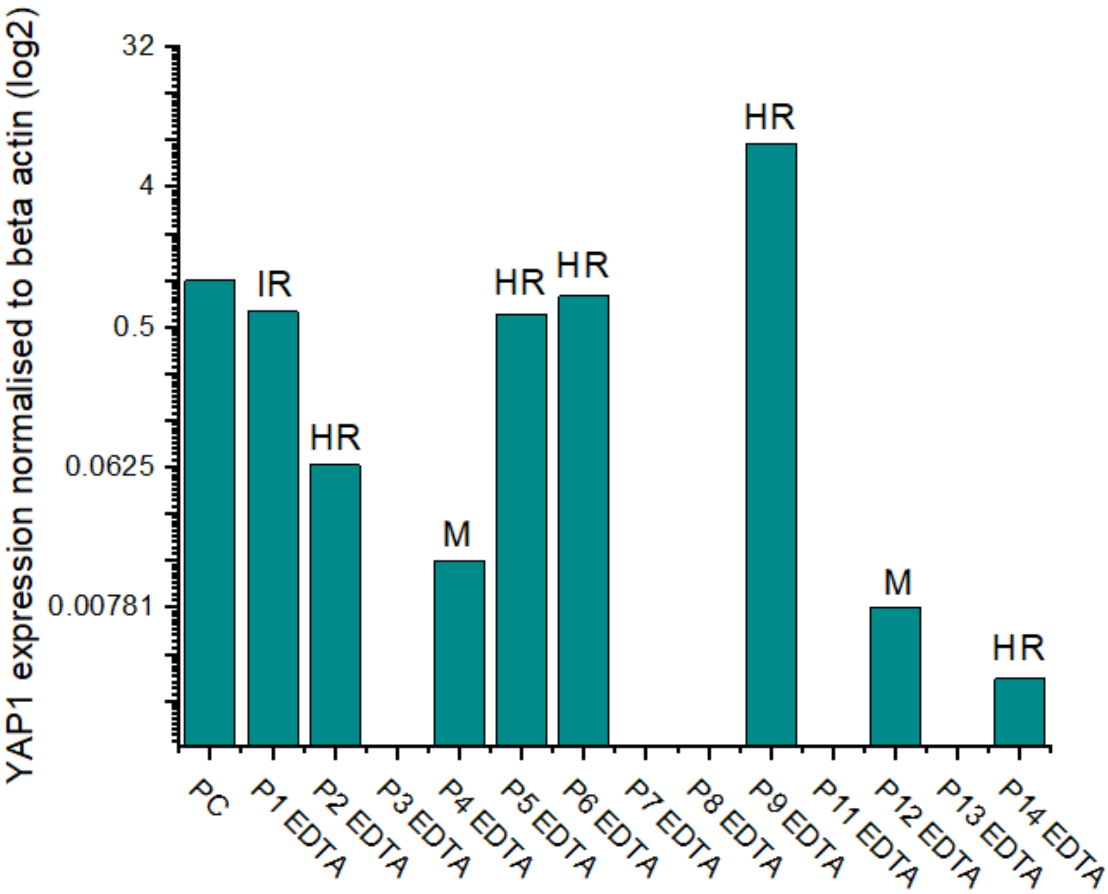

Figure S9: The relative YAP1 expression to beta actin in clinical samples using the YAP1 RT-qPCR primer set 1 and the ΔΔCt method. Sample YAP1 expression was relative to the beta actin expression of the individual sample and normalised to the positive control of 0.5 ng of LNCaP cDNA. HR = high risk, IR = intermediate risk, m = metastatic.

### 10 AR-V7 mRNA RT-qPCR and RT-pHLAMP benchtop detection and PSA levels.

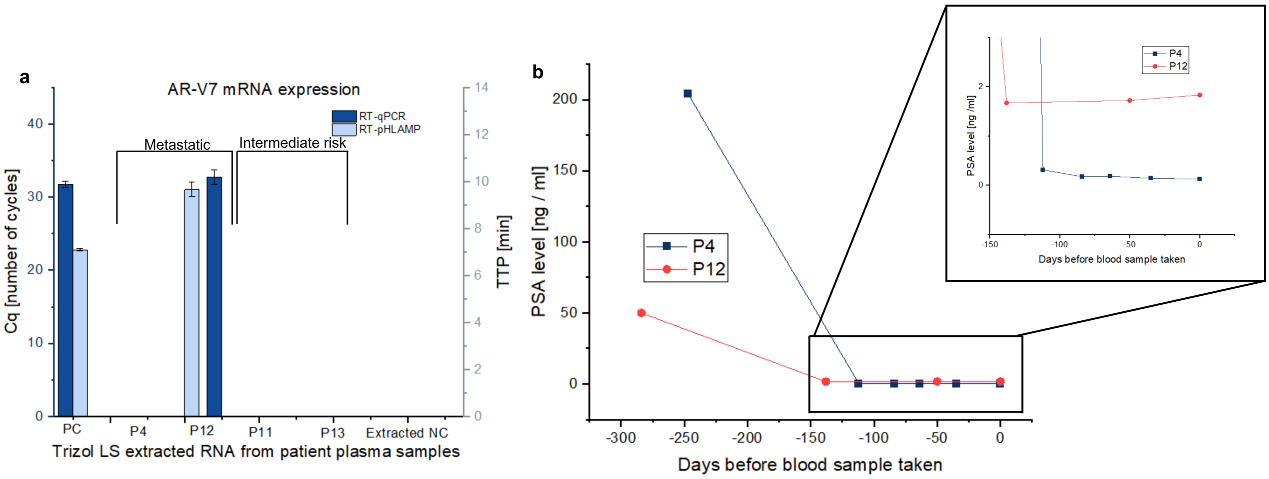

Figure S10: **(a)**: The detection of AR-V7 with both RT-qPCR and RT-pHLAMP benchtop assays in metastatic patients (P4 and P12) and intermediate risk patients (P11 and P13). **(b)**: The PSA levels of P4 and P12 from previous blood tests leading up to blood sample collection.
